# Social and spatial disparities in heat-related mortality in Italy: a nationwide small-area study

**DOI:** 10.64898/2026.07.06.26357399

**Authors:** Barbara Sodano, Connor Gascoigne, Di Xi, Xinyi Chen, Francesca de’Donato, Paolo Vineis, Garyfallos Konstantinoudis

**Author notes:** Corresponding author: Garyfallos Konstantinoudis, Grantham Institute for Climate Change and the Environment, Imperial College London, London, UK *Email address* (Garyfallos Konstantinoudis).

## Abstract

**Background:** Spatial variation in heat-related mortality remains poorly understood, particularly at fine geographical scales. We conducted a nationwide small-area study to examine the association between spatial variation in heat-related mortality and environmental, demographic, health, and socio-economic factors.

**Methods:** We obtained daily all-cause mortality data for people aged ≥ 65 years during the summers of 2011– 2023 and linked them with municipality-level daily temperature estimates from the ERA5-Land reanalysis dataset. We applied a two-stage Bayesian hierarchical model to estimate small-area heat-related mortality and assess the contribution of community characteristics to spatial variability.

**Findings:** Heat-related mortality showed marked geographical differences, with the highest rates in southern and southeastern Italy. Across municipalities, the relative risk at the 90th temperature percentile, relative to the minimum mortality temperature, ranged from 1.06 to 1.33. The heat-attributable fraction exceeded 6% in several southern municipalities, while excess mortality surpassed 8 deaths per 1,000 inhabitants in parts of the Po Valley, Tuscany, Apulia, and Sicily. National heat-attributable mortality peaked in 2022, with an estimated 17,828 deaths (95% credible intervals: 17,339, 18,285) among older adults. Municipalities with higher average temperatures, less green space, higher obesity prevalence, and more residents aged ≥ 85 years had higher heat-related mortality. Educational attainment and employment were among the strongest modifiers of spatial variation.

**Interpretation:** Our findings highlight substantial small-area differences in heat-related mortality across Italy and identify socio-economic deprivation as a key determinant of vulnerability. Heat is likely to disproportionately affect disadvantaged communities, reinforcing the need for adaptation strategies addressing social inequality.

**Funding:** Imperial College Research Fellowship; Italian Ministry of Health PNC (CUP J55I22004450001); NIHR Imperial Biomedical Research Center (BRC NIHR203323).

**Research in context:** *Evidence before this study:* Previous studies have shown that heat increases mortality risk, especially among older adults, and that the burden is unevenly distributed across Europe and within Italy. However, evidence on the community characteristics explaining fine-scale spatial disparities in heat-related mortality remains limited.

*Added value of this study:* This nationwide small-area study estimates municipality-specific heat-related mortality across 7,895 Italian municipalities from 2011 to 2023 and links spatial variation in risk to environmental, demographic, health, and socio-economic factors.

*Implications of all the available evidence:* Heat adaptation strategies should account for local, social and environmental vulnerability, with targeted interventions for disadvantaged communities and areas with high population vulnerability.

## Introduction

Exposure to extreme ambient heat is a major driver of excess mortality and morbidity, especially among older adults and individuals with pre-existing chronic conditions ^1,2^. With ongoing climate change, heat-related health risks are becoming an increasingly important public health concern across Europe. Recent summers have caused substantial heat-related mortality, for example, summer 2025 temperatures were estimated to cause approximately 25,000 deaths across 854 European cities ^3^. This burden was unevenly distributed, with the highest heat-related mortality observed in Mediterranean cities ^3^. Likewise, a previous study of summer 2022 identified substantial small-area variation in vulnerability across Mediterranean countries, including Italy ^4^. Evidence from Italy has documented marked geographical heterogeneity in heat-related mortality across cities and regions, together with temporal changes and modification by environmental factors such as air pollution ^5,6^. However, the factors driving this fine-scale spatial differences remain poorly understood.

Estimates of mortality due to high temperatures exhibit geographical disparities in many contexts ^7^. Those spatial disparities may be due to environmental, demographic and socio-economic characteristics ^8,9^. Higher levels of urbanisation have been associated with increased heatwave-related mortality risk ^10^, whereas higher levels of green spaces appear to mitigate the adverse health effects of high temperatures ^11^. Socio-economic position has also been suggested as a potential determinant of vulnerability to heat exposure, with higher heat-related mortality values observed among individuals with lower socio-economic position ^12^. However, spatial inequalities may also reflect unidentified contextual factors.

In Italy, several studies have documented pronounced spatial disparities in the association between ambient temperature and mortality. Regional analyses assessing the effects of both extreme heat and cold have reported significant differences in temperature-mortality relationships across Italian regions, indicating that the mortality burden associated with non-optimal temperatures varies geographically ^5^. Furthermore, evidence of heterogeneity has also been observed at finer spatial scales (municipalities). In central-eastern Italy, a multi-city analysis of heat-wave effects on elderly mortality found substantial spatial disparities between cities ^13^. More recently, a case time-series study conducted across Italian municipalities confirmed strong small-area variability in the temperature–mortality association ^14^. Despite this consistent evidence, little is known about the community factors that may underlie such heterogeneity, limiting a more complete understanding of disparities in vulnerability to high summer temperatures across Italy.

In this nationwide analysis of heat-related mortality in Italy, we investigated the association between heat exposure and daily all-cause mortality across 7,895 municipalities from 2011 to 2023. We used a two-stage modelling approach to estimate municipality-specific heat–mortality associations and then assess the contribution of community factors to the observed spatial disparities. From the first-stage model, we estimated municipality-specific relative risks, attributable fractions and excess deaths. In the second-stage, we assessed the contribution of environmental, demographic, health and socio-economic factors to these spatial disparities.

## Methods

### Study design and participants

In this nationwide small-area study in Italy during 2011-2023, we retrieved daily all-cause mortality and the resident population data for 7,895 municipalities from the Italian National Institute of Statistics (ISTAT; https://www.istat.it/). As we are focusing on the impact of heat, we restricted the analysis on the summer months (June, July and August). We also restricted the analyses to individuals aged 65 years or older, as this group is the most vulnerable to heat-related mortality and morbidity ^4^. Population data was also retrieved from ISTAT and was available on an annual basis from January 1st of each year (Online Supplement Text S1). We reported this study in accordance with the Strengthening the Reporting of Observational Studies in Epidemiology (STROBE) guidelines, the completed checklist is provided in Online Supplement Table S1.

### Exposure

We retrieved mean daily temperature data during 2011-2023 from the Copernicus Climate Change Service ERA5-Land reanalysis product (https://cds.climate.copernicus.eu). We linked temperature to mortality records by matching municipality centroids to the corresponding ERA5-Land grid cells (9 km x 9 km). Consistent with evidence that the effects of heat on mortality are acute, we defined exposure as the average temperature on the day of death and the three preceding days (lag 0-3) ^15^.

### Confounders

We adjusted for national holidays, year, seasonality (as measured by day-of-the-year), and day-of-the-week. We also accounted for relative humidity (lag 0-3), estimated using the ERA-5 reanalysis data and linking it with the outcome as we did for temperature.

### Community factors

To evaluate heterogeneity in heat-related mortality we considered community factors representing environmental conditions (e.g., the Normalized Difference Vegetation Index, and average temperature from ERA-5), demographic characteristics (e.g., population density, urbanicity, proportion of individuals older than 85 years), health indicators (e.g., number of hospital beds per population, health expenditure per population, and smoking and obesity prevalence), and socio-economic deprivation (e.g., the municipality-level 2018 Fragility Index). In a secondary analysis, we examined the associations between heat-related risks and each of the 12 component indicators of the fragility index. These included environmental and territorial factors, such as transport-related emissions, unsorted municipal waste per capita, protected natural areas, landslide risk exposure, land consumption, and accessibility to essential services, as well as socio-demographic and economic factors, including the dependency index, low educational attainment, employment rate among those aged 20–64, migration rate, firm density, and employment in low-productivity sectors. For a detailed description of the sources and hypotheses behind the inclusion of the selected community factors see Online Supplement Text S2. Definitions of the Fragility Index components are provided in Online Supplement Table S2.

### Statistical analysis

We employed a two-staged Bayesian hierarchical model. In the first-stage, we estimated spatial differences by fitting a model that allows for municipality-level non-linear associations between temperature and mortality ^16^. In the second-stage, we examined the contribution of the above described community factors to the estimated spatial disparities in heat-related mortality.

Let *Y*_*dtm*_ be the number of deaths on the *d*^th^ day, in the *t*^th^ year in the *m*^th^ municipality, where *m* = 1, …, 7895, *d* = 1, …, 92, and *t* = 1, …, 13, respectively; and *P*_*dtm*_ the population size. We modelled *Y*_*dtm*_ using the following Poisson likelihood and linear predictor,

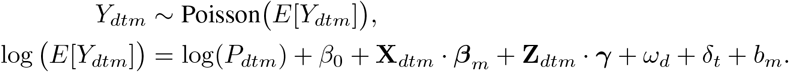

where *β*_0_ is the global intercept, **X**_*dtm*_ represents the basis matrix of mean temperature (lags 0–3) with municipality-specific regression coefficients 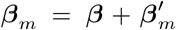 ^16,17^. To define the basis matrix we used natural cubic splines, with knots placed at the 10th, 75th, and 90th percentiles of the municipality-specific temperature distribution. The matrix **Z**_*dtm*_ accounts for day of the week and public holidays and relative humidity with ***γ*** the respective vector of regression coefficients. The final three terms, *ω*_*d*_, *δ*_*t*_, and *b*_*m*_ account for any unmeasured, residual confounding across the seasonality, year, and municipality domains. For more information about the specification of the priors see Online Supplement Text S3.

In the second stage of the analysis, we examined the influence of the selected spatial effect modifiers on the heat-vulnerabilities: excess deaths due to heat (ERH), the attributable fraction due to heat (AFH), and the relative risk (RR) at the 90th percentile of temperature. For more details on the calculation of these metrics, see Online Supplement Text S4. Focusing on ERH, we fitted three different models assuming a normal distribution with mean *μ*_*m*_ and variance 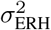:

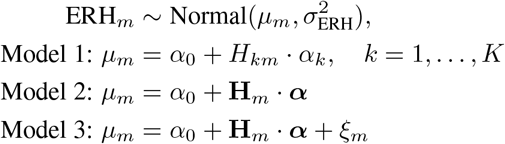

where *α*_0_ is a global intercept, *α*_*k*_ is the regression coefficient associated with the *k*-th spatial effect modifier *H*_*km*_, and ***α*** is the vector of regression coefficients for all modifiers collected in **H**_*m*_. The term *ξ*_*m*_ captures residual local spatial correlation through an intrinsic conditional auto-regressive (ICAR) prior ^18^. For the fixed effects *α*_0_ and ***α***, we specified flat, non-informative priors. These three models are designed to complementary assess aspects of spatial heat-related disparities: Model 1 (univariable model) examines the marginal contribution of each community factor individually; Model 2 (multivariable models) assesses the joint contribution of all selected spatial effect modifiers; and Model 3 (spatial) extends Model 2 by additionally accounting for residual spatial autocorrelation not captured by the measured modifiers.

All analyses were conducted in R version 4.4.1 (R Core Team, 2024), and all models were fitted using the Integrated Nested Laplace Approximation (INLA) ^19^. In the first stage, we report the posterior median of heat vulnerability for each municipality and the posterior probability that it exceeds the national average. In the second stage, we report posterior medians and 95% credible intervals (CrI) for ***α*** from Models 1–3. To account for uncertainty in the first-stage estimates, we fitted each second-stage model to 200 draws from the posterior distribution of ERH_*m*_ and pooled the resulting estimates. The analysis code is available in the project GitHub repository.

### Sensitivity analysis

In a first sensitivity analysis, we allowed seasonality and long-term trends in the first-stage model to vary across municipalities. This specification makes our approach more comparable to previous studies that fitted models separately for each city or region ^4^. In a second sensitivity analysis, we removed all years following the start of the COVID-19 pandemic in Italy, 2020-2023. For both the first and second sensitivity analyses, we compared the national and municipality effect of temperature to the main analysis including those years. In a third sensitivity analysis, we defined heat vulnerability using the posterior distribution of the relative risk at the 95th and 99th temperature percentiles, and compared the resulting spatial patterns with those obtained using the 90th temperature percentile.

## Results

### Exploratory analysis

Between 2011 and 2023, there were in total 1,746,943 deaths among older adults during summers 2011-2023 in Italy (Table 1). The daily mortality rate among older adults exhibited short-term variability and a slight upward trend over time (Online Supplement Figure S1, left panel). The spatial distribution of mortality rates (Online Supplement Figure S1, right panel) appeared relatively homogeneous across municipalities, with no strong or consistent geographic pattern. A clear increasing trend in mean daily summer temperature is observed over the study period (Online Supplement Figure S2, left panel). Spatially, mean summer temperature (Online Supplement Figure S2, right panel) displayed a clear geographical pattern, with higher values in southern and lowland areas and lower temperatures in northern and mountainous regions. The spatial distribution of the included covariates is presented in the Online Supplement Figures S3-S4.

**Table 1.** Annual attributable numbers (AN), attributable fraction of heat (AFH), population and deaths among individuals aged ≥65 years in Italy, 2011–2023.

| Year | Population | Deaths | AN (median, 95% CrI) | AFH (median, 95% CrI) |
| --- | --- | --- | --- | --- |
| 2011 | 12,254,516 | 124,232 | 4,252 (4,110–4,382) | 2.78% (2.70%–2.88%) |
| 2012 | 12,519,205 | 128,272 | 9,841 (9,526–10,139) | 5.47% (5.30%–5.67%) |
| 2013 | 12,741,455 | 127,568 | 5,259 (5,054–5,444) | 3.25% (3.15%–3.38%) |
| 2014 | 12,979,066 | 123,144 | 1,173 (1,076–1,267) | 1.30% (1.25%–1.35%) |
| 2015 | 13,181,761 | 139,192 | 11,773 (11,472–12,059) | 5.41% (5.24%–5.59%) |
| 2016 | 13,321,182 | 127,927 | 4,346 (4,167–4,503) | 2.80% (2.70%–2.90%) |
| 2017 | 13,461,945 | 135,319 | 12,454 (12,115–12,771) | 6.03% (5.85%–6.22%) |
| 2018 | 13,565,148 | 130,658 | 6,801 (6,566–7,020) | 3.52% (3.40%–3.66%) |
| 2019 | 13,693,215 | 135,512 | 10,425 (10,082–10,737) | 5.13% (4.96%–5.31%) |
| 2020 | 13,859,090 | 136,737 | 6,770 (6,543–6,993) | 3.78% (3.65%–3.92%) |
| 2021 | 13,941,531 | 144,398 | 11,324 (11,011–11,616) | 5.59% (5.42%–5.76%) |
| 2022 | 14,051,404 | 156,673 | 17,828 (17,339–18,285) | 7.10% (6.87%–7.34%) |
| 2023 | 14,181,297 | 137,311 | 12,542 (12,278–12,807) | 6.00% (5.83%–6.18%) |
| TOTAL | 176,750,815 | 1,746,943 | 114,812 (111,549–117,856) | 4.55% (4.42%–4.71%) |

### Temperature-mortality association

The effect of heat on all-cause mortality was J-shaped across all municipalities, with steep increases for temperatures above 80^th^ percentile, Figure 1 and Online Supplement Figure S5. The annual median number of deaths attributable to heat nationwide ranged from 1,172 deaths in 2014 (95% CrI: 1,075, 1,266) to 17,828 deaths in 2022 (95% CrI: 17,339, 18,285). Although 2023 showed a reduction compared with 2022, attributable mortality remained high (12,542 deaths; 95% CrI: 12,278, 12,807). Region-level attributable numbers (AN) of deaths due to non-optimal summer temperatures (posterior medians and 95% CrIs) are reported in the online Supplement Material (Supplement Table S3).

**Figure 1.**
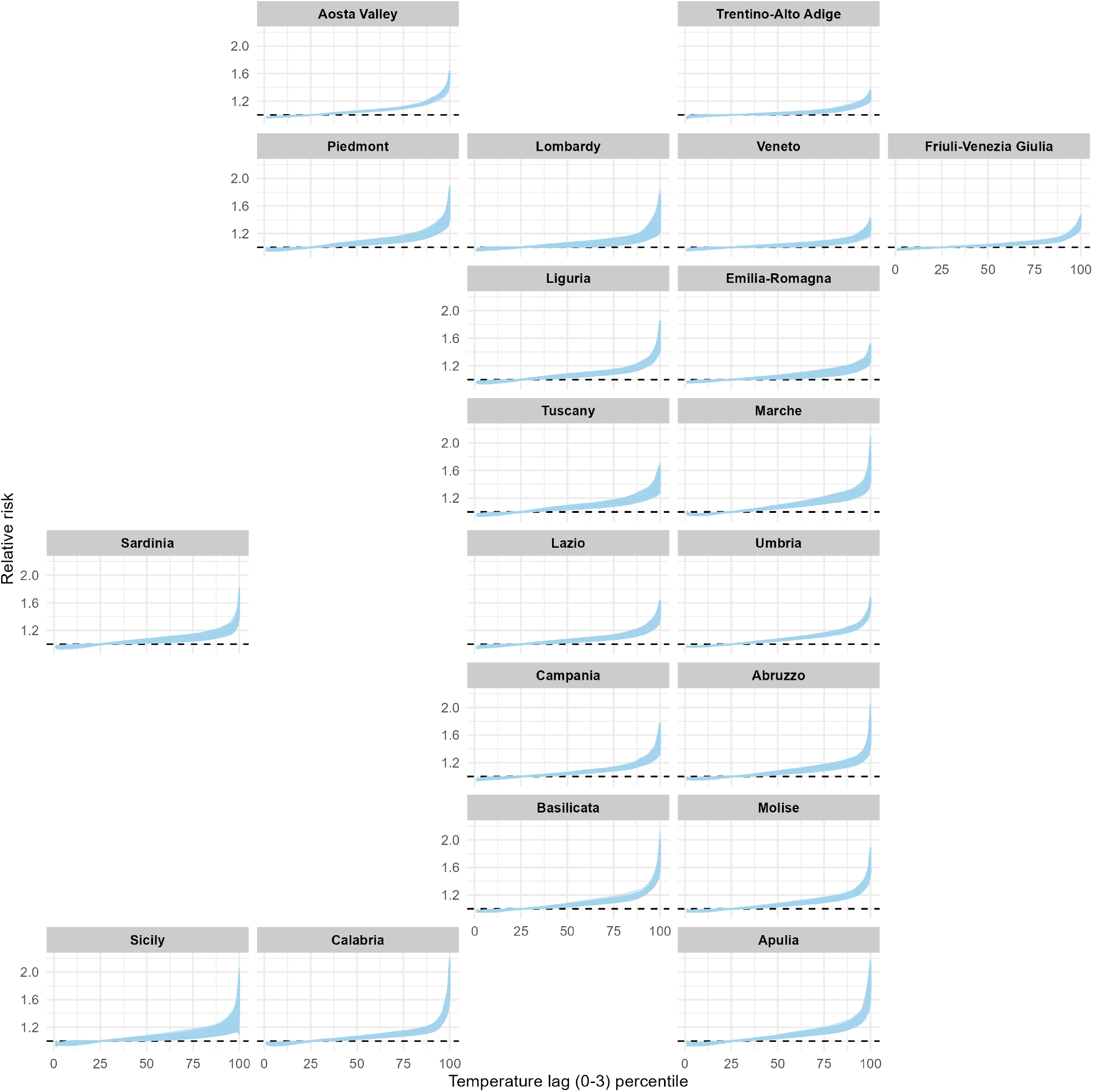
Median posterior of relative mortality risk at temperature percentiles for each municipality (blue lines) at the 20 Italian regions

### Spatial disparities in heat-related risks

Figure 2 shows spatial heat vulnerability across municipalities. Panels A and D represent the relative mortality risk at the 90th temperature percentile and the posterior probability that the RR is higher than the national average. The RR varies from 1.06 to 1.33 between Italian municipalities, with values higher than 1.22 in Piedmont, Lombardy, Liguria, Tuscany, Marche and Apulia. Panels B and E show the proportion of deaths attributed to heat and the posterior probability that this is larger than the nationwide average. In most municipalities in North-East, the median attributable fraction of heat (AFH) ranged between 0% and 2%, while the AFH was higher than 6% in many municipalities in the south. We observed more than 8 deaths per thousand population attributable to summer heat exposure in many municipalities of the Po Valley, Tuscany, Marche, Apulia and Sicily, Panel C in Figure 2. In the North-East regions the ERH ranges from 0 to 3 deaths per thousand population, Panel C in Figure 2.

**Figure 2.**
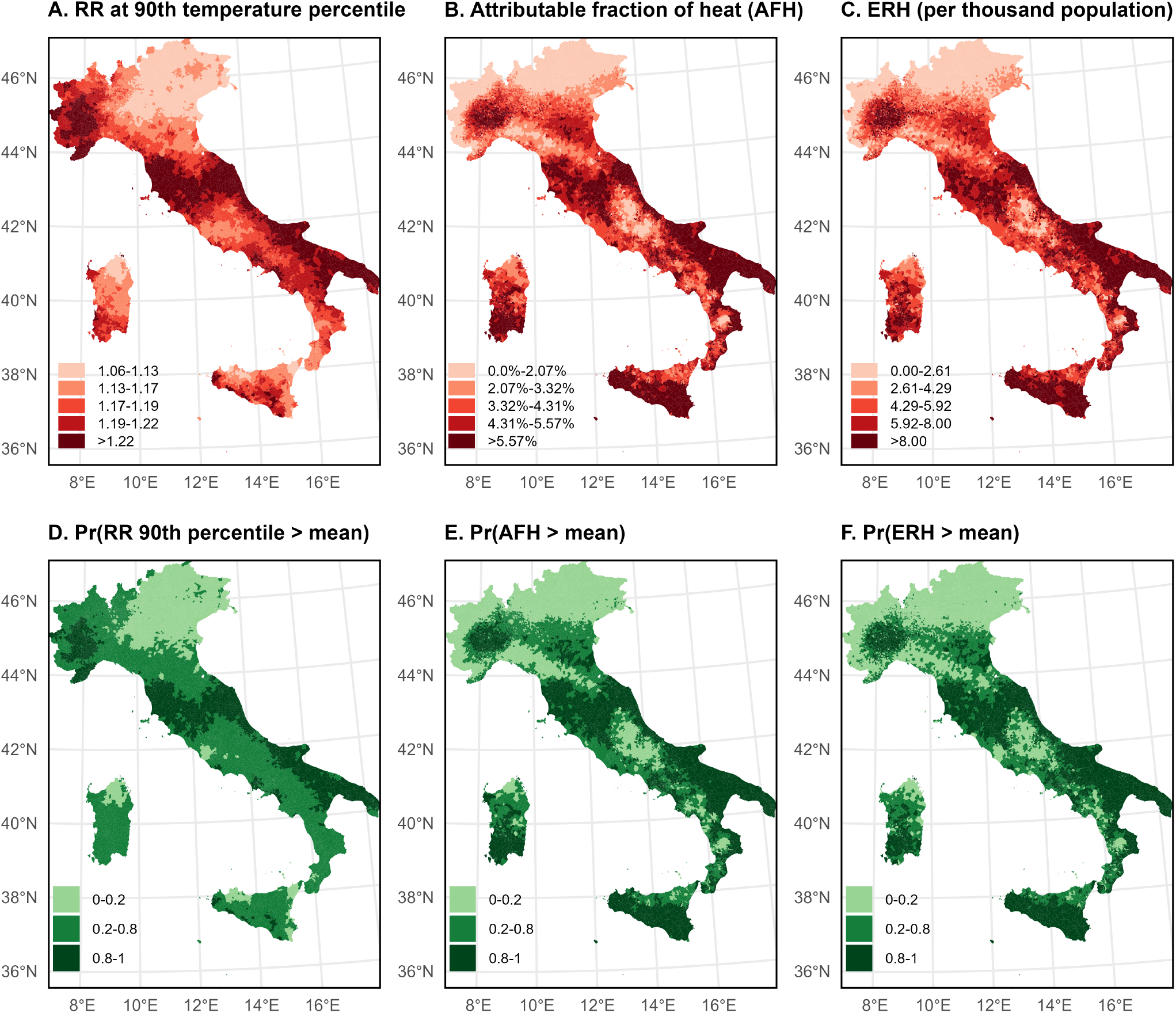
A. RR of all-cause mortality at 90th temperature percentile at municipality level; B. Attributable Fraction of heat (AFH) in each municipality; C. Excess mortality rates (ERH) per thousand population attributable to heat in each municipality; D. The exceedance probability of RR at 90th temperature percentile in the area is higher than the mean value of the RR at 90th percentile across the country; E. The exceedance probability of attributable fraction of heat (AFH) in the area is higher than the mean value of AFH across the country; F. The exceedance probability of excess mortality rates (ERH) attributable to heat in the area is higher than the mean value of the ERH across the country.

### Community factors contribution

Figure 3 shows the posterior medians and 95% CrIs for the association with the selected community factors. The standard deviations of the selected continuous effect modifiers are reported in the Online Supplement Material (Supplement Table S4). In the multivariable spatial model (Model 3), the clearest associations were observed for environmental conditions and socio-economic deprivation. Municipalities with higher average temperatures had higher heat-related mortality, with estimates that were consistent in direction and magnitude across all three model specifications. By contrast, greater green space coverage was strongly associated with lower heat-related mortality: each standard deviation increase in green space was associated with about 4.4 fewer heat-related deaths per 10,000 population (95% CrI: −5.58, −3.44; Online Supplement Table S5). The fragility index showed a clear dose–response pattern, with higher levels of the index associated with greater heat-related mortality. This pattern was consistent across models, although attenuated after spatial adjustment. In Model 3, compared with fragility level 1, fragility level 5, corresponding to the highest level of fragility, was associated with an increase of 3.10 heat-related deaths per 10,000 population (95% CrI: 0.82, 5.91) (Figure 3 and Online Supplement Table S5). Lower levels of fragility, such as levels 2–4, showed weaker associations. We found weaker evidence of an association with high proportion of people older than 85 years and higher prevalence of obesity. Evidence for the rest of the community factors was inconclusive. These findings were broadly consistent when we considered the RR at the 90th temperature percentile and the AFH, Online Supplement Figures S6-S7 and Tables S6-S7.

**Figure 3.**
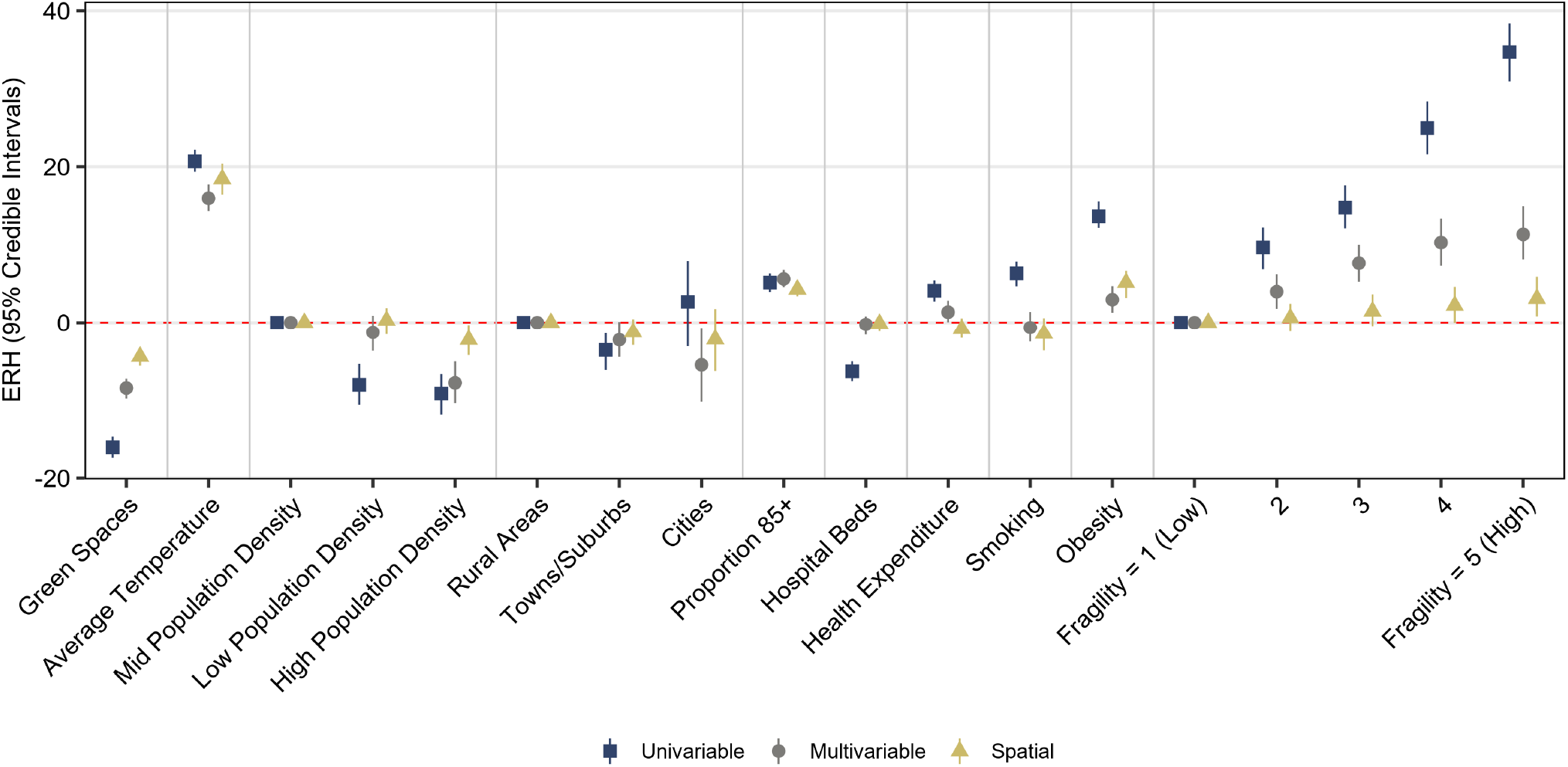
Effect of covariates on heat-related excess mortality rate after uncertainty propagation (the point represents the median estimate of each coefficient and the error bars show the 95% CrI of coefficients).

To identify which dimensions of fragility underpinned these associations, we examined the individual components of the Fragility Index (Figure 4 and Online Supplement Table S8). The positive association with the overall Fragility Index appears to be primarily driven by socio-economic indicators, namely employment rate, low educational attainment, the proportion of workers in low-productivity sectors, and firms per capita. Transport emissions were also positively associated with heat vulnerability (Figure 4).

**Figure 4.**
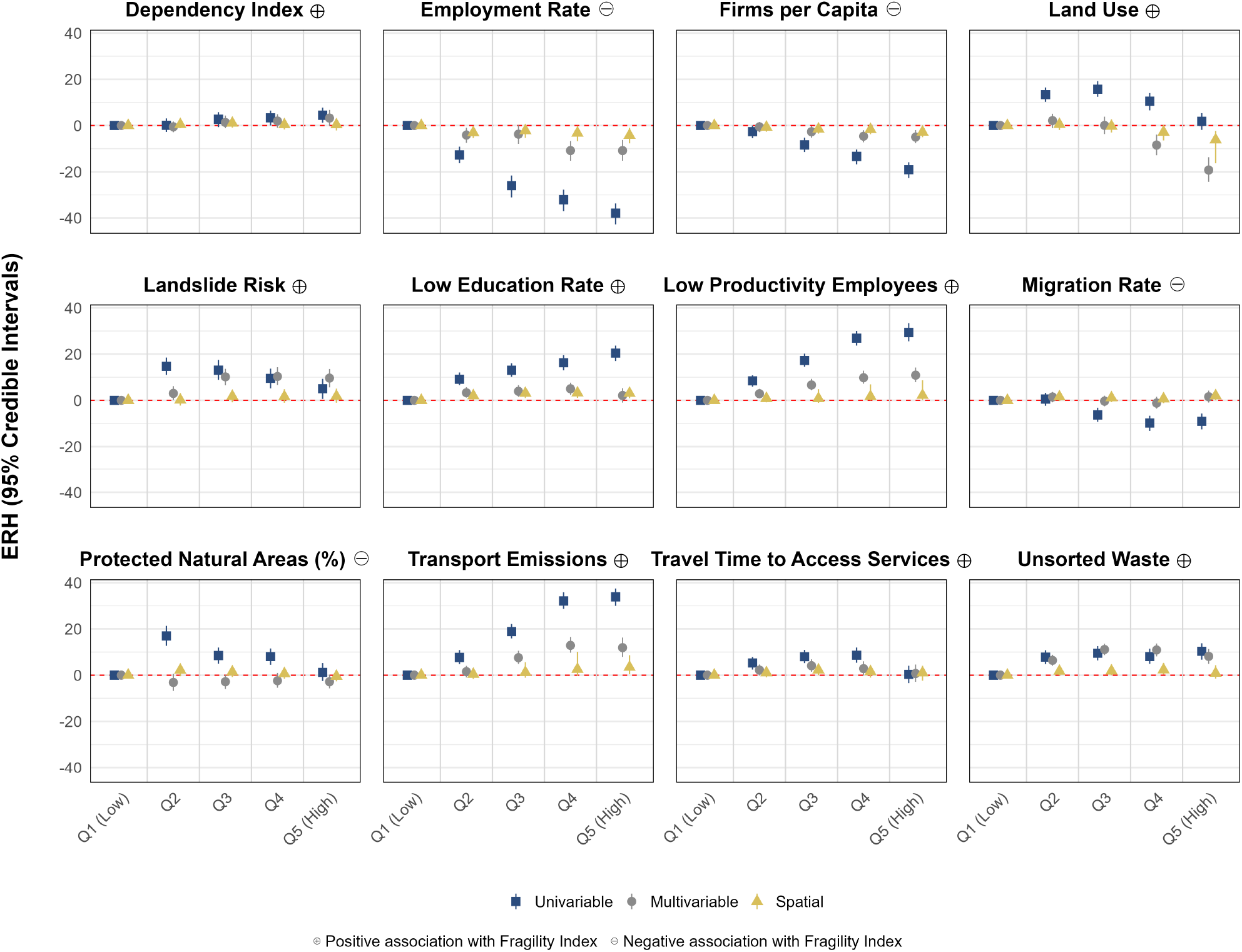
Effect of the single components of the Fragility Index on heat-related excess mortality rate after uncertainty propagation (the point represents the median estimate of each coefficient and the error bars show the 95% CrI of coefficients).

### Sensitivity analysis

The sensitivity analysis including the interaction terms and removing the years after the start of the COVID-19 pandemic in Italy showed no significant differences when comparing against the main analysis (Online Supplement, Figures S8 and S9). Likewise, the spatial patterns of relative risk at the 95th and 99th temperature percentiles were broadly consistent with those based on the 90th temperature percentile (Online Supplement, Figure S10).

## Discussion

This study provides the first comprehensive assessment of municipal heat-related mortality disparities in Italy, considering environmental, demographic, and socio-economic factors associated with it. Our findings reveal pronounced geographical disparities, with the greatest vulnerability in southern and southeastern Italy. Most notably, we found a strong socio-economic gradient in heat vulnerability: municipal fragility showed a clear dose–response association driven primarily by socio-economic indicators. Heat vulnerability was higher in municipalities with higher average temperatures, higher obesity prevalence, more residents aged ≥85 years, and less green space.

This study has several strengths. First, it provides a nationwide assessment of heat-related mortality at a very fine spatial scale. Second, the use of a Bayesian hierarchical modelling framework with spatial structure allows for robust estimation of municipality-specific temperature–mortality relationships while accounting for spatial dependence across neighbouring municipalities. Moreover, the Bayesian hierarchical model allows for uncertainty to be accounted for and fully propagated when aggregating results to higher spatial resolution and in the follow-up second-stage analysis.

This study has several limitations. Although both stages adjusted for multiple confounders, residual confounding cannot be excluded, particularly from unmeasured factors such as housing conditions or access to cooling. In addition, several contextual covariates, including obesity, smoking prevalence, and deprivation, were only available for a single time point and may not capture temporal changes or within-municipality heterogeneity over the study period. Moreover, smoking and obesity estimates were available only at the regional level for individuals aged 14 years and older, and may not accurately represent these risk factors among adults aged ≥65 years. Although inclusion of spatial autocorrelation partly addressed geographic structure, residual spatial confounding or overadjustment remains possible, as suggested by the attenuation of some associations after inclusion of spatial random effects. Outdoor temperature was used as a proxy and may not reflect personal heat exposure. Finally, the small-area design may still be affected by ecological bias, although likely to a lesser extent given the fine spatial scale.

Our results are consistent with previous evidence from Italy. The J-shaped temperature–mortality relationship matches earlier nationwide and city-specific studies ^20,21^, and our estimates of heat-attributable deaths among adults aged 65 years or older in 2022 (17,828) and 2023 (12,542) are close to those reported in recent European analyses ^4,22,23^. The spatial pattern we identified is consistent with previous evidence from Italy, which has shown higher heat-related mortality in the south and lower impacts in the north-east ^14,24^.

We found that certain environmental characteristics were associated with excess heat-related mortality. In particular, municipalities with higher average temperatures had greater heat-related mortality, likely reflecting greater exposure to thermal stress. We also found that greater green space coverage was associated with lower heat vulnerability, consistent with previous modelling and epidemiological studies showing protective effects of greenness ^11,25,26^. The protective effect of green space may reflect cooler microclimates and co-benefits including reduced air pollution and improved physical, social and mental well-being ^27^.

Municipalities with a higher proportion of residents aged aged ≥85 years tended to have greater heat-related mortality, consistent with evidence that the oldest age groups are particularly vulnerable to heat because of impaired thermoregulation, reduced physiological reserve, and a higher prevalence of chronic conditions ^28^. By contrast, we found limited evidence that urbanicity or population density were important predictors after adjustment. Previous studies have reported heterogeneous findings regarding the role of urbanicity in heat-related mortality. For example, Sera et al. ^29^ observed higher heat-related mortality in more densely populated urban areas, while other studies found no evidence of differences according to population density, despite observing associations with measures of urbanicity ^30^. A recent nationwide French study reported higher heat-related mortality in urban than rural communities, partly explained by NO_2_ pollution and urban heat islands ^31^.

Of the health indicators examined, obesity showed the clearest positive association with heat-related mortality. This is biologically plausible because excess body mass impairs thermoregulation, reduces heat dissipation and increases cardiovascular strain ^32^. Obesity may also reflect poorer underlying health through its association with chronic conditions that increase heat vulnerability. By contrast, we found limited evidence of associations with hospital beds or health expenditure. In line with recent European evidence, we found that hospital beds and health expenditure are not consistently associated with heat-related vulnerability ^12^. This may reflect the availability of these indicators only at provincial level, limiting their ability to capture local differences in healthcare access. In addition, the distribution of healthcare resources, including hospital beds, is shaped by broader urban–rural differences and service organization, which may not directly translate into differences in heat-related vulnerability at the local level.

Socio-economic deprivation emerged as one of the strongest predictors of spatial heat vulnerability. Municipal fragility showed an association with heat-related mortality, reinforcing the critical role of social and economic inequalities as heat-related mortality risk factors. This is consistent with previous evidence linking heat-related mortality to social disadvantage ^29,33^. Higher levels of deprivation have also been associated with greater heat-related vulnerability across European regions, with socio-economic disadvantage contributing to the overall temperature-related mortality burden ^12^. Populations living in more deprived areas may have limited access to adaptive resources, such as air conditioning, adequate housing, and healthcare, and may also face a higher burden of pre-existing disease, all of which can increase heat-related mortality. However, previous Italian studies conducted at broader spatial scales have reported inconsistent findings regarding the role of socio-economic conditions and educational level in heat-related mortality. The nationwide coverage and much finer spatial resolution of our analysis may have allowed us to better capture small-area socio-economic disparities across municipalities.

In conclusion, this study provides a detailed assessment of heat-related mortality across 7,895 municipalities in Italy and shows that its spatial distribution is shaped by both environmental exposures and social vulnerabilities. The marked spatial heterogeneity in heat-related mortality across Italian municipalities indicates that adaptation cannot rely on uniform strategies, but instead requires geographically targeted action at a fine spatial scale. The populations most affected by heat are also the least able to protect themselves because socially disadvantaged communities face greater exposure, fewer environmental protections and reduced access to housing, cooling, healthcare and green space. In this way, extreme heat reinforces existing inequalities, underscoring the importance of integrating environmental and social vulnerability into climate policy to reduce the unequal health impacts of heat in a warming climate.

## Data availability statement

The data used in this study is open access. Temperature data: https://cds.climate.copernicus.eu/, mortality data: https://demo.istat.it/#sezione1., population data: https://demo.istat.it/#sezione1., fragility index and components, obesity and smoking prevalence: https://esploradati.istat.it/databrowser/#/it/dw/categories, health expenditure and number of hospital beds: https://www.dati.salute.gov.it/ and urbanicity: https://ec.europa.eu/eurostat/web/ cities/database

## Code availability

Code for performing the analysis is provided in the [project GitHub repository](https://github.com/bsodano-98/Italy-Heat-Community-Factors).

## Ethics

The study is about secondary, aggregate anonymised data so no ethical permission is required.

## Acknowledgments

G.K. is supported by an Imperial College Research Fellowship.

B.S. is supported by the Italian Ministry of Health – PNC “HEALTH AND EQUITY CO-BENEFITS IN SUPPORT OF CLIMATE CHANGE RESPONSE PLANS IN ITALY” (CUP J55I22004450001).

All authors acknowledge Infrastructure support for the Department of Epidemiology and Biostatistics provided by the NIHR Imperial Biomedical Research Centre (BRC NIHR203323).

B.S. gratefully acknowledges Prof. Marta Blangiardo for providing access to the course on Bayesian Reasoning and Methods for Spatio-Temporal Data, which contributed to the development of this work.

## Contributors

G.K. conceived the study; G.K. and P.V. supervised the work. B.S. developed the initial study protocol in discussion with D.X., X.C., P.V. and G.K.. B.S. downloaded and cleaned all the population, mortality, exposure and community factors data. B.S. developed the first version of the first-stage statistical model and C.G. validated, revised and ran the analysis. B.S. developed the second-stage models, which was revised and validated by G.K.. B.S. wrote the first version of the manuscript. All authors read and approved the final manuscript.

## Declaration of interests

The authors declare no competing interests.

## Notes

### Competing Interest Statement

The authors have declared no competing interest.

### Author Declarations

The study is about secondary, aggregate anonymised data so no ethical permission is required. The data used in this study is open access. Links to access mortality and population data: https://demo.istat.it/#sezione1.

