## Supplementary Material for "Social and spatial disparities in heat-related mortality in Italy: a nationwide small-area study"

S10 A. Median of the posterior distribution of the relative mortality risk at 95th temperature percentile at municipality level; B.Median of the posterior distribution of the relative mortality risk at 99th temperature percentile at municipality level; C. The exceedance probability of relative mortality risk at 95th temperature percentile in the area is higher than the mean value of the RR at 95th percentile across the country; D. The exceedance probability of relative mortality risk at 99th temperature percentile in the area is higher than the mean value of the RR at 95th percentile across the country 27

**Table S1:** STROBE Statement checklist of items that should be included in reports of observational studies.

| Section/topic | Item No. | Recommendation | Page No. |
|---|---|---|---|
| **Title and abstract** | | | |
| Title and abstract | 1(a) | Indicate the study's design with a commonly used term in the title or the abstract | 1 |
| | 1(b) | Provide in the abstract an informative and balanced summary of what was done and what was found | 1 |
| **Introduction** | | | |
| Background/rationale | 2 | Explain the scientific background and rationale for the investigation being reported | 1–2 |
| Objectives | 3 | State specific objectives, including any prespecified hypotheses | 2 |
| **Methods** | | | |
| Study design | 4 | Present key elements of study design early in the paper | 2 |
| Setting | 5 | Describe the setting, locations, and relevant dates, including periods of recruitment, exposure, follow-up, and data collection | 2–3 |
| Participants | 6(a) | Cohort study: give the eligibility criteria, and the sources and methods of selection of participants; describe methods of follow-up. Case-control study: give the eligibility criteria, and the sources and methods of case ascertainment and control selection; give the rationale for the choice of cases and controls. Cross-sectional study: give the eligibility criteria, and the sources and methods of selection of participants | 2–3 |
| | 6(b) | Cohort study: for matched studies, give matching criteria and number of exposed and unexposed. Case-control study: for matched studies, give matching criteria and the number of controls per case | Not applicable |
| Variables | 7 | Clearly define all outcomes, exposures, predictors, potential confounders, and effect modifiers. Give diagnostic criteria, if applicable | 2–4 |
| Data sources/measurement | 8* | For each variable of interest, give sources of data and details of methods of assessment or measurement. Describe comparability of assessment methods if there is more than one group | 2–3 |
| Bias | 9 | Describe any efforts to address potential sources of bias | 3–4 |
| Study size | 10 | Explain how the study size was arrived at | 2–3 |
| Quantitative variables | 11 | Explain how quantitative variables were handled in the analyses. If applicable, describe which groupings were chosen and why | 2–4 |
| Statistical methods | 12(a) | Describe all statistical methods, including those used to control for confounding | 3–4 |
| | 12(b) | Describe any methods used to examine subgroups and interactions | 4 |

*Continued on next page*

*Table S1 continued from previous page*

| Section/topic | Item No. | Recommendation | Page No. |
|---|---|---|---|
| | 12(c) | Explain how missing data were addressed | Not applicable |
| | 12(d) | Cohort study: if applicable, explain how loss to follow-up was addressed. Case-control study: if applicable, explain how matching of cases and controls was addressed. Cross-sectional study: if applicable, describe analytical methods taking account of sampling strategy | Not applicable |
| | 12(e) | Describe any sensitivity analyses | 4 |
| **Results** | | | |
| Participants | 13(a)* | Report numbers of individuals at each stage of study, for example numbers potentially eligible, examined for eligibility, confirmed eligible, included in the study, completing follow-up, and analysed | 4 |
| | 13(b) | Give reasons for non-participation at each stage | Not applicable |
| | 13(c) | Consider use of a flow diagram | Not applicable |
| Descriptive data | 14(a)* | Give characteristics of study participants, for example demographic, clinical, and social characteristics, and information on exposures and potential confounders | 4–5 |
| | 14(b) | Indicate number of participants with missing data for each variable of interest | Not applicable |
| | 14(c) | Cohort study: summarise follow-up time, for example average and total amount | 4–5 |
| Outcome data | 15* | Cohort study: report numbers of outcome events or summary measures over time. Case-control study: report numbers in each exposure category, or summary measures of exposure. Cross-sectional study: report numbers of outcome events or summary measures | 4–5 |
| Main results | 16(a) | Give unadjusted estimates and, if applicable, confounder-adjusted estimates and their precision, for example 95% confidence interval. Make clear which confounders were adjusted for and why they were included | 4–5 |
| | 16(b) | Report category boundaries when continuous variables were categorized | 4–5 |
| | 16(c) | If relevant, consider translating estimates of relative risk into absolute risk for a meaningful time period | Not applicable |
| Other analyses | 17 | Report other analyses done, for example analyses of subgroups and interactions, and sensitivity analyses | 5 |
| **Discussion** | | | |
| Key results | 18 | Summarise key results with reference to study objectives | 5 |
| Limitations | 19 | Discuss limitations of the study, taking into account sources of potential bias or imprecision. Discuss both direction and magnitude of any potential bias | 6 |

*Continued on next page*

*Table S1 continued from previous page*

| **Section/topic** | **Item No.** | **Recommendation** | **Page No.** |
|---|---|---|---|
| Interpretation | 20 | Give a cautious overall interpretation of results considering objectives, limitations, multiplicity of analyses, results from similar studies, and other relevant evidence | 6 |
| Generalisability | 21 | Discuss the generalisability, or external validity, of the study results | 6–7 |
| **Other information** | | | |
| Funding | 22 | Give the source of funding and the role of the funders for the present study and, if applicable, for the original study on which the present article is based | 7 |

*Give information separately for cases and controls in case-control studies and, if applicable, for exposed and unexposed groups in cohort and cross-sectional studies.

Note: An Explanation and Elaboration article discusses each checklist item and gives methodological background and published examples of transparent reporting. The STROBE checklist is best used in conjunction with this article, freely available on the websites of PLOS Medicine at http://www.plosmedicine.org/, Annals of Internal Medicine at http://www.annals.org/, and Epidemiology at http://www.epidem.com/. Information on the STROBE Initiative is available at https://www.strobe-statement.org/.

**Table S2:** Components of the Municipal Fragility Index (IFC, ISTAT 2018), calculated at the municipal level and referring to year 2018 data.

| **Indicator** | **Definition** | **Association with fragility** |
|---|---|---|
| Dependency Index | Percentage ratio between the population aged 0–19 years and ≥65 years and the population aged 20–64 years. The indicator corresponds to the sum of youth and old-age dependency indices and measures the demographic burden on the working-age population. Data were derived from the Permanent Population and Housing Census. Unit of measure: percentage values. | Positive |
| Employment Rate | Percentage ratio between employed individuals aged 20–64 years and the resident population aged 20–64 years. The indicator measures labour market participation among the working-age population. Data were derived from the Permanent Population and Housing Census. Unit of measure: percentage values. | Negative |
| Firms per Capita | Ratio between the stock of active local business units and the resident population (per 1,000 inhabitants). The indicator measures the territorial density of economic activities and local productive capacity. Data were derived from ISTAT processing of the Asia Local Units register and the Permanent Population and Housing Census. Unit of measure: ventile classes (1 = minimum; 20 = maximum). | Negative |
| Land Consumption | Percentage of artificial land cover related to urban settlement dynamics over the total municipal area. Data derive from ISPRA land consumption monitoring and quantify anthropogenic land transformation. Unit of measure: percentage values. | Positive |
| Landslide Risk | Percentage of municipal territorial surface classified as high or very high landslide risk (P3–P4 classes of the Hydrogeological Asset Plan). The indicator captures exposure to severe hydrogeological instability using ISPRA and ISTAT territorial data. Reference years differed across IFC editions (2017 for IFC 2018). Unit of measure: percentage values. | Positive |
| Low Education Rate | Percentage of residents aged 25–64 years with educational attainment not exceeding lower secondary school or vocational initiation qualifications. The indicator measures the prevalence of low educational attainment within the adult population. Data were derived from the Permanent Population and Housing Census. Unit of measure: percentage values. | Positive |
| Low Productivity Employees | Percentage of employees working in local units below the first quartile of the nominal labour productivity distribution within industry and service sectors (ATECO 2007 classification). The indicator measures the concentration of workers employed in low-productivity sectors. In the absence of local units, municipalities were imputed to the first ventile. Data were derived from Frame-SBS territorial statistics. Unit of measure: ventile classes (1 = minimum; 20 = maximum). | Positive |
| Migration Rate | Ratio between net migration balance and the initial resident population (per 1,000 inhabitants). Positive values indicate population attraction, whereas negative values indicate population loss. Net migration was estimated using the residual method based on demographic variation. Data were derived from demographic balance statistics and the Permanent Population and Housing Census. Unit of measure: per 1,000 inhabitants. | Negative |
| Protected Natural Areas (%) | Percentage of municipal territory covered by protected terrestrial natural areas included in the EUAP list or Natura 2000 Network (SCI/ZPS/ZSC). The indicator measures the presence of protected environmental resources within the municipality. Data were derived from ISPRA and the Ministry for the Environment and Energy Security. Unit of measure: percentage values. | Negative |
| Transport Emissions | Number of circulating high-emission passenger vehicles (Euro 0–3 categories) per 100 inhabitants. The indicator measures the prevalence of highly polluting vehicles within the municipality. Data were derived from ACI and ISTAT population statistics. Unit of measure: per 100 inhabitants. | Positive |
| Travel Time to Access Services | Average travel time required to reach the nearest service hub, including hospitals, secondary schools, and railway stations. The indicator measures accessibility to essential services and infrastructure using commercial road network and territorial databases. For the IFC 2018 edition, the reference year for service accessibility data was 2019. Unit of measure: minutes. | Positive |
| Unsorted Waste | Per-capita amount of unsorted municipal solid waste (kg per inhabitant). The indicator measures the burden of non-separated urban waste production. Data were derived from ISPRA waste registers and ISTAT population statistics. Unit of measure: kilograms per inhabitant. | Positive |

Notes: All indicators refer to the year 2018 and were calculated at the municipal level (Italian municipalities/comuni). Positive association indicates that higher values correspond to higher municipal fragility, whereas negative association indicates that higher values correspond to lower fragility. Data and metadata are publicly available from the Italian National Institute of Statistics (ISTAT): https://esploradati.istat.it/databrowser/#/it/dw/categories/IT1,Z0930TER,1.0/CFI_MUN/IT1,DF_COMP_FRA_IND_MUNICIPAL_01,1.0.

**Figure S1:** Mean daily mortality rate among people over 65 years old during summers from 2011 to 2023 (left panel); Spatial pattern of the mortality rate among people over 65 years old across Italian municipalities (right panel).

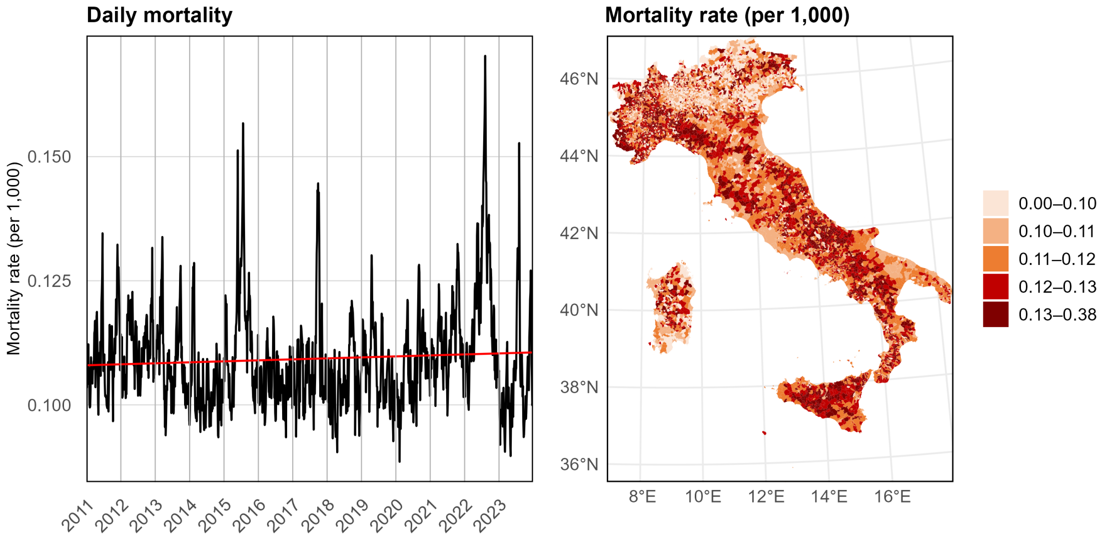

**Figure S2:** Daily mean temperature during summers from 2011 to 2023 (left panel); Spatial pattern of the mean temperature exposure across Italian municipalities (right panel).

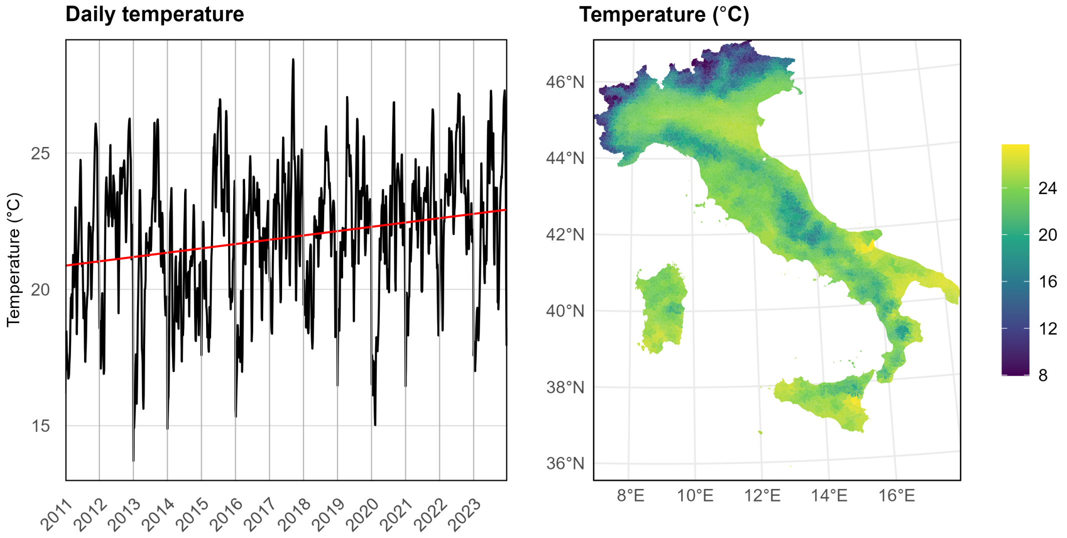

**Figure S3:** Values of effect modifiers in Italy at municipal level (Average Temperature, Green Spaces, Population Density, Urbanicity, Proportion 85+, Fragility), at provincial level (Hospital Beds by population, Health Expenditure by population) and at regional level (Obesity and Smoking).

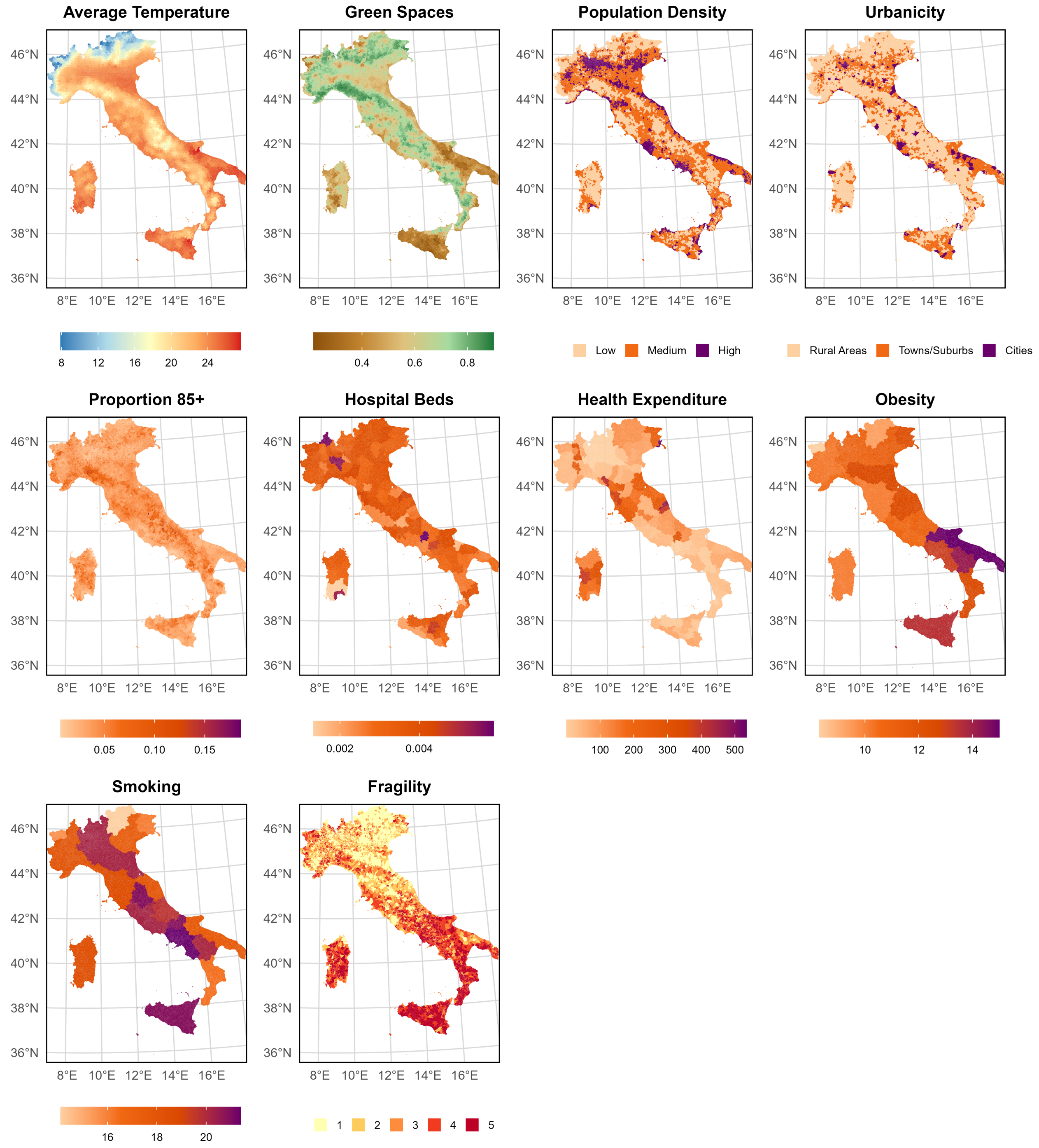

**Figure S4:** Values of the 12 components of the Fragility Index in Italy at municipal level (quintiles)

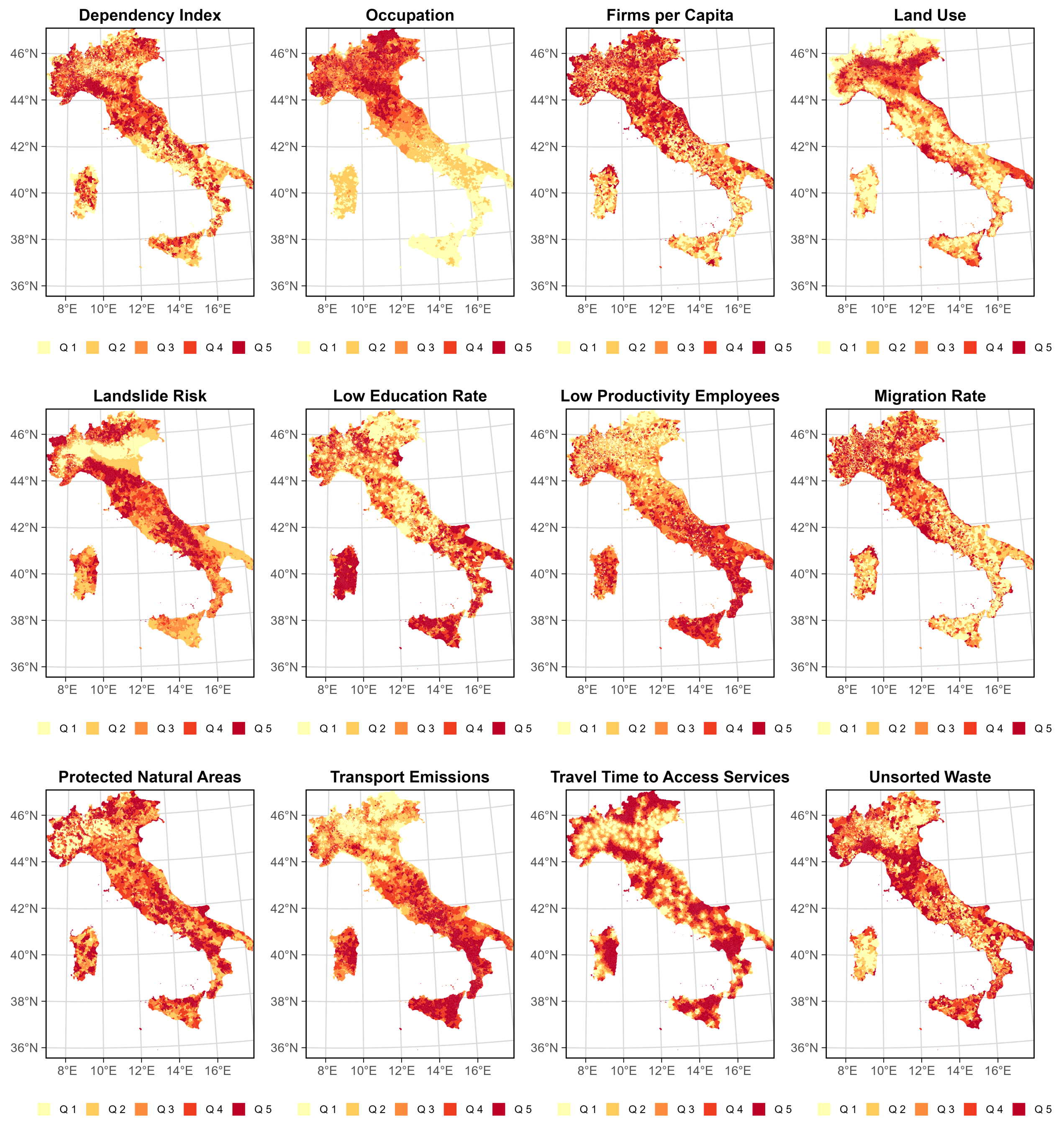

**Figure S5:** Median of the posterior distribution of the relative mortality risk across the 7,895 municipalities during 2011-2023 by temperature percentiles. The green line represents the nationwide relative mortality risk calculated as the median of the municipality-specific median relative mortality risk per percentile

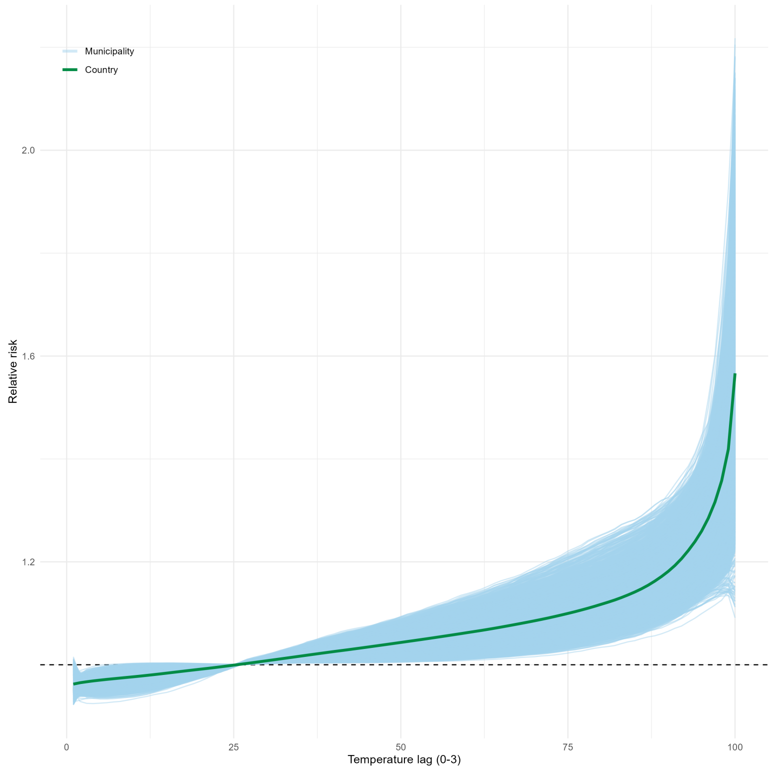

**Table S3:** Region-level attributable number (AN) of deaths due to non-optimal summer temperatures (median and 95% CrI).

| Region | AN (median, 95% CrI) |
|---|---|
| Piedmont | 12,126 (11,444–12,771) |
| Liguria | 4,140 (3,772–4,495) |
| Lombardy | 15,240 (14,329–16,203) |
| Veneto | 5,121 (4,550–5,618) |
| Lazio | 8,961 (8,261–9,613) |
| Abruzzo | 3,283 (3,027–3,566) |
| Aosta Valley | 278 (219–338) |
| Trentino-Alto Adige/South Tyrol | 916 (752–1,103) |
| Campania | 9,716 (9,040–10,374) |
| Sicily | 9,728 (9,272–10,152) |
| Sardinia | 2,909 (2,724–3,098) |
| Friuli Venezia Giulia | 1,837 (1,535–2,145) |
| Emilia-Romagna | 7,654 (6,990–8,252) |
| Marche | 4,904 (4,562–5,218) |
| Tuscany | 9,413 (8,794–9,961) |
| Umbria | 2,416 (2,206–2,652) |
| Apulia | 10,006 (9,384–10,592) |
| Molise | 821 (707–926) |
| Basilicata | 1,380 (1,238–1,539) |
| Calabria | 3,997 (3,620–4,361) |

**Table S4:** Standard deviations of the selected continuous effect modifiers

| Variable | Standard Deviation | Units |
|---|---|---|
| Average Temperature | 3.12 | °C |
| Green Spaces | 0.14 | NDVI index |
| Proportion 85+ | 0.0169 | % |
| Hospital Beds | 0.00076 | number of hospital beds/population |
| Health Expenditure | 95.92 | health expenditure/population |
| Obesity | 1.41 | % |
| Smoking | 1.74 | % |

**Table S5:** Association estimates of ERH from univariable, multivariable, and spatial effect modification models

| Variable | Model | Mean | Median | 95% CrI lower | 95% CrI upper |
|---|---|---|---|---|---|
| **Univariable model** | | | | | |
| Green Spaces | Univariable | **−16.03** | **−16.04** | **−17.38** | **−14.66** |
| Average Temperature | Univariable | **20.71** | **20.68** | **19.34** | **22.18** |
| Hospital Beds | Univariable | **−6.27** | **−6.32** | **−7.57** | **−5.03** |
| Mid Population Density (ref.) | Univariable | 0.00 | 0.00 | 0.00 | 0.00 |
| Low Population Density | Univariable | **−8.00** | **−8.04** | **−10.60** | **−5.36** |
| High Population Density | Univariable | **−9.19** | **−9.17** | **−11.87** | **−6.64** |
| Proportion 85+ | Univariable | **5.14** | **5.15** | **3.95** | **6.35** |
| Health Expenditure | Univariable | **4.08** | **4.11** | **2.71** | **5.44** |
| Smoking | Univariable | **6.30** | **6.34** | **4.68** | **7.85** |
| Obesity | Univariable | **13.71** | **13.66** | **12.16** | **15.55** |
| Rural Areas (ref.) | Univariable | 0.00 | 0.00 | 0.00 | 0.00 |
| Towns/Suburbs | Univariable | **−3.63** | **−3.55** | **−6.15** | **−1.38** |
| Cities | Univariable | 2.68 | 2.67 | −3.08 | 7.89 |
| Fragility = 1 (Low) | Univariable | 0.00 | 0.00 | 0.00 | 0.00 |
| Fragility = 2 | Univariable | **9.63** | **9.66** | **6.87** | **12.21** |
| Fragility = 3 | Univariable | **14.77** | **14.76** | **12.08** | **17.62** |
| Fragility = 4 | Univariable | **24.97** | **24.95** | **21.57** | **28.38** |
| Fragility = 5 (High) | Univariable | **34.63** | **34.69** | **30.93** | **38.36** |
| **Multivariable model** | | | | | |
| Green Spaces | Multivariable | **−8.47** | **−8.46** | **−9.80** | **−7.27** |
| Average Temperature | Multivariable | **16.01** | **15.95** | **14.30** | **17.75** |
| Hospital Beds | Multivariable | −0.29 | −0.24 | −1.58 | 0.80 |
| Mid Population Density (ref.) | Multivariable | 0.00 | 0.00 | 0.00 | 0.00 |
| Low Population Density | Multivariable | −1.33 | −1.28 | −3.65 | 0.90 |
| High Population Density | Multivariable | **−7.79** | **−7.79** | **−10.41** | **−5.03** |
| Proportion 85+ | Multivariable | **5.63** | **5.62** | **4.56** | **6.76** |
| Health Expenditure | Multivariable | **1.36** | **1.35** | **0.05** | **2.84** |
| Smoking | Multivariable | −0.62 | −0.65 | −2.45 | 1.40 |
| Obesity | Multivariable | **2.99** | **2.96** | **1.29** | **4.71** |
| Rural Areas (ref.) | Multivariable | 0.00 | 0.00 | 0.00 | 0.00 |
| Towns/Suburbs | Multivariable | −2.21 | −2.25 | −4.43 | 0.09 |
| Cities | Multivariable | **−5.40** | **−5.47** | **−10.20** | **−0.71** |
| Fragility = 1 (Low) | Multivariable | 0.00 | 0.00 | 0.00 | 0.00 |
| Fragility = 2 | Multivariable | **4.00** | **3.99** | **1.76** | **6.23** |
| Fragility = 3 | Multivariable | **7.62** | **7.65** | **5.24** | **10.00** |
| Fragility = 4 | Multivariable | **10.32** | **10.28** | **7.33** | **13.34** |
| Fragility = 5 (High) | Multivariable | **11.40** | **11.33** | **8.13** | **14.95** |
| **Spatial model** | | | | | |
| Green Spaces | Spatial | **−4.44** | **−4.41** | **−5.58** | **−3.44** |
| Average Temperature | Spatial | **18.44** | **18.44** | **16.42** | **20.41** |
| Hospital Beds | Spatial | −0.13 | −0.12 | −1.14 | 0.88 |
| Mid Population Density (ref.) | Spatial | 0.00 | 0.00 | 0.00 | 0.00 |
| Low Population Density | Spatial | 0.28 | 0.29 | −1.52 | 1.85 |
| High Population Density | Spatial | **−2.30** | **−2.22** | **−4.23** | **−0.33** |
| Proportion 85+ | Spatial | **4.30** | **4.28** | **3.38** | **5.27** |
| Health Expenditure | Spatial | −0.79 | −0.84 | −2.01 | 0.55 |

| Variable | Model | Mean | Median | 95% CrI lower | 95% CrI upper |
|---|---|---|---|---|---|
| Smoking | Spatial | −1.43 | −1.43 | −3.60 | 0.57 |
| Obesity | Spatial | **5.09** | **5.13** | **3.18** | **6.66** |
| Rural Areas (ref.) | Spatial | 0.00 | 0.00 | 0.00 | 0.00 |
| Towns/Suburbs | Spatial | −1.25 | −1.27 | −2.91 | 0.43 |
| Cities | Spatial | −2.17 | −2.17 | −6.25 | 1.73 |
| Fragility = 1 (Low) | Spatial | 0.00 | 0.00 | 0.00 | 0.00 |
| Fragility = 2 | Spatial | 0.58 | 0.61 | −1.15 | 2.44 |
| Fragility = 3 | Spatial | 1.51 | 1.47 | −0.47 | 3.65 |
| Fragility = 4 | Spatial | 2.22 | 2.21 | −0.04 | 4.60 |
| Fragility = 5 (High) | Spatial | **3.17** | **3.10** | **0.82** | **5.91** |

**Figure S6:** Effect of covariates on attributable fraction of heat after uncertainty propagation (the point represents the median estimate of each coefficient and the error bars show the 95% CrI of coefficients).

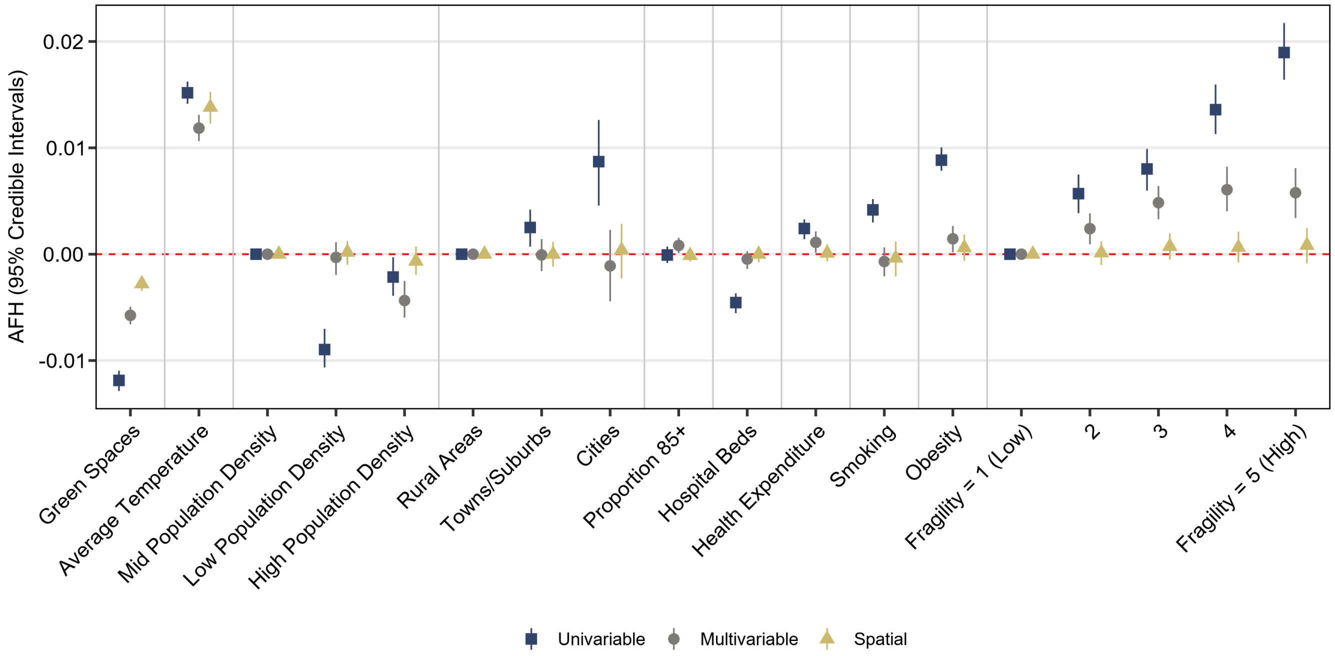

**Figure S7:** Effect of covariates on mortality relative risk at 90th temperature percentile after uncertainty propagation (the point represents the median estimate of each coefficient and the error bars show the 95% CrI of coefficients).

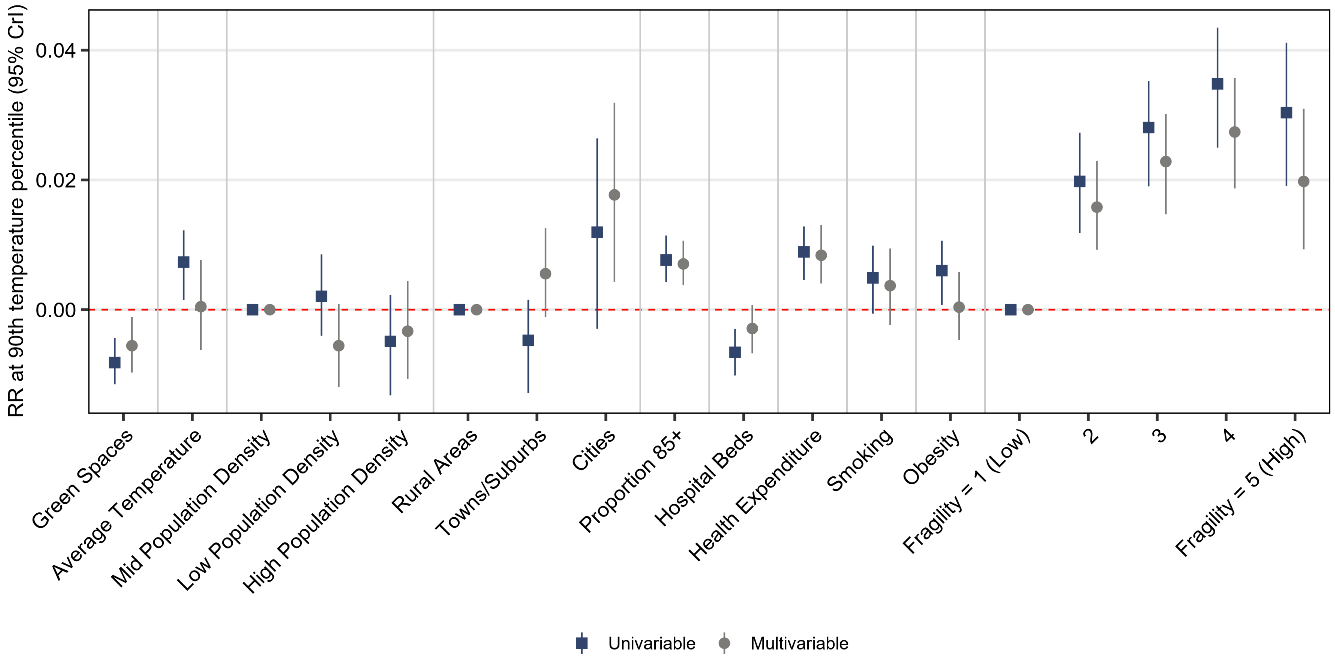

**Table S6:** Association estimates of AFH from univariable, multivariable, and spatial effect modification models

| Variable | Model | Mean | Median | 95% CrI lower | 95% CrI upper |
|---|---|---|---|---|---|
| **Univariable model** | | | | | |
| Green Spaces | Univariable | **−0.011 88** | **−0.011 87** | **−0.012 86** | **−0.010 96** |
| Average Temperature | Univariable | **0.015 17** | **0.015 17** | **0.014 14** | **0.016 23** |
| Hospital Beds | Univariable | **−0.004 56** | **−0.004 55** | **−0.005 55** | **−0.003 68** |
| Mid Population Density (ref.) | Univariable | 0.000 00 | 0.000 00 | 0.000 00 | 0.000 00 |
| Low Population Density | Univariable | **−0.008 94** | **−0.008 97** | **−0.010 65** | **−0.007 02** |
| High Population Density | Univariable | **−0.002 14** | **−0.002 15** | **−0.003 91** | **−0.000 29** |
| Proportion 85+ | Univariable | −0.000 08 | −0.000 08 | −0.000 82 | 0.000 72 |
| Health Expenditure | Univariable | **0.002 39** | **0.002 41** | **0.001 41** | **0.003 27** |
| Smoking | Univariable | **0.004 15** | **0.004 17** | **0.002 98** | **0.005 19** |
| Obesity | Univariable | **0.008 89** | **0.008 84** | **0.007 85** | **0.010 03** |
| Rural Areas (ref.) | Univariable | 0.000 00 | 0.000 00 | 0.000 00 | 0.000 00 |
| Towns/Suburbs | Univariable | **0.002 49** | **0.002 51** | **0.000 71** | **0.004 19** |
| Cities | Univariable | **0.008 72** | **0.008 70** | **0.004 57** | **0.012 62** |
| Fragility = 1 (Low) | Univariable | 0.000 00 | 0.000 00 | 0.000 00 | 0.000 00 |
| Fragility = 2 | Univariable | **0.005 69** | **0.005 69** | **0.003 86** | **0.007 49** |
| Fragility = 3 | Univariable | **0.007 97** | **0.008 01** | **0.005 98** | **0.009 89** |
| Fragility = 4 | Univariable | **0.013 58** | **0.013 59** | **0.011 29** | **0.015 95** |
| Fragility = 5 (High) | Univariable | **0.018 98** | **0.018 96** | **0.016 40** | **0.021 75** |
| **Multivariable model** | | | | | |
| Green Spaces | Multivariable | **−0.005 75** | **−0.005 75** | **−0.006 60** | **−0.004 94** |
| Average Temperature | Multivariable | **0.011 88** | **0.011 85** | **0.010 64** | **0.013 10** |
| Hospital Beds | Multivariable | −0.000 47 | −0.000 46 | −0.001 38 | 0.000 28 |

| Variable | Model | Mean | Median | 95% CrI lower | 95% CrI upper |
|---|---|---|---|---|---|
| Mid Population Density (ref.) | Multivariable | 0.000 00 | 0.000 00 | 0.000 00 | 0.000 00 |
| Low Population Density | Multivariable | −0.000 35 | −0.000 30 | −0.001 94 | 0.001 13 |
| High Population Density | Multivariable | **−0.004 32** | **−0.004 36** | **−0.005 95** | **−0.002 51** |
| Proportion 85+ | Multivariable | **0.000 83** | **0.000 82** | **0.000 11** | **0.001 54** |
| Health Expenditure | Multivariable | **0.001 10** | **0.001 11** | **0.000 14** | **0.002 15** |
| Smoking | Multivariable | −0.000 68 | −0.000 69 | −0.002 08 | 0.000 65 |
| Obesity | Multivariable | **0.001 48** | **0.001 45** | **0.000 18** | **0.002 66** |
| Rural Areas (ref.) | Multivariable | 0.000 00 | 0.000 00 | 0.000 00 | 0.000 00 |
| Towns/Suburbs | Multivariable | −0.000 09 | −0.000 07 | −0.001 58 | 0.001 43 |
| Cities | Multivariable | −0.001 07 | −0.001 10 | −0.004 43 | 0.002 28 |
| Fragility = 1 (Low) | Multivariable | 0.000 00 | 0.000 00 | 0.000 00 | 0.000 00 |
| Fragility = 2 | Multivariable | **0.002 40** | **0.002 40** | **0.000 93** | **0.003 84** |
| Fragility = 3 | Multivariable | **0.004 85** | **0.004 84** | **0.003 29** | **0.006 41** |
| Fragility = 4 | Multivariable | **0.006 12** | **0.006 06** | **0.004 03** | **0.008 23** |
| Fragility = 5 (High) | Multivariable | **0.005 77** | **0.005 78** | **0.003 41** | **0.008 09** |
| **Spatial model** | | | | | |
| Green Spaces | Spatial | **−0.002 79** | **−0.002 79** | **−0.003 46** | **−0.002 06** |
| Average Temperature | Spatial | **0.013 80** | **0.013 82** | **0.012 28** | **0.015 26** |
| Hospital Beds | Spatial | 0.000 00 | −0.000 01 | −0.000 74 | 0.000 70 |
| Mid Population Density (ref.) | Spatial | 0.000 00 | 0.000 00 | 0.000 00 | 0.000 00 |
| Low Population Density | Spatial | 0.000 15 | 0.000 18 | −0.000 99 | 0.001 24 |
| High Population Density | Spatial | −0.000 65 | −0.000 66 | −0.001 93 | 0.000 71 |
| Proportion 85+ | Spatial | −0.000 12 | −0.000 12 | −0.000 68 | 0.000 39 |
| Health Expenditure | Spatial | 0.000 10 | 0.000 10 | −0.000 68 | 0.000 93 |
| Smoking | Spatial | −0.000 39 | −0.000 37 | −0.002 09 | 0.001 18 |
| Obesity | Spatial | 0.000 62 | 0.000 61 | −0.000 62 | 0.001 85 |
| Rural Areas (ref.) | Spatial | 0.000 00 | 0.000 00 | 0.000 00 | 0.000 00 |
| Towns/Suburbs | Spatial | −0.000 02 | −0.000 01 | −0.001 18 | 0.001 16 |
| Cities | Spatial | 0.000 38 | 0.000 39 | −0.002 27 | 0.002 86 |
| Fragility = 1 (Low) | Spatial | 0.000 00 | 0.000 00 | 0.000 00 | 0.000 00 |
| Fragility = 2 | Spatial | 0.000 13 | 0.000 12 | −0.001 01 | 0.001 21 |
| Fragility = 3 | Spatial | 0.000 74 | 0.000 72 | −0.000 50 | 0.001 96 |
| Fragility = 4 | Spatial | 0.000 65 | 0.000 64 | −0.000 77 | 0.002 12 |
| Fragility = 5 (High) | Spatial | 0.000 81 | 0.000 83 | −0.000 86 | 0.002 46 |

**Table S7:** Association estimates of RR at 90th temperature percentile from univariable and multivariable models

| Variable | Model | Mean | Median | 95% CrI lower | 95% CrI upper |
|---|---|---|---|---|---|
| **Univariable model** | | | | | |
| Green Spaces | Univariable | **−0.008 12** | **−0.008 16** | **−0.011 49** | **−0.004 38** |
| Average Temperature | Univariable | **0.007 31** | **0.007 34** | **0.001 51** | **0.012 23** |
| Hospital Beds | Univariable | **−0.006 56** | **−0.006 58** | **−0.010 14** | **−0.002 95** |
| Mid Population Density (ref.) | Univariable | 0.000 00 | 0.000 00 | 0.000 00 | 0.000 00 |
| Low Population Density | Univariable | 0.002 10 | 0.002 06 | −0.004 01 | 0.008 52 |
| High Population Density | Univariable | −0.005 08 | −0.004 89 | −0.013 21 | 0.002 29 |
| Proportion 85+ | Univariable | **0.007 71** | **0.007 66** | **0.004 26** | **0.011 43** |
| Health Expenditure | Univariable | **0.008 88** | **0.008 91** | **0.004 59** | **0.012 83** |
| Smoking | Univariable | 0.004 86 | 0.004 90 | −0.000 61 | 0.009 87 |
| Obesity | Univariable | **0.005 97** | **0.006 02** | **0.000 71** | **0.010 64** |
| Rural Areas (ref.) | Univariable | 0.000 00 | 0.000 00 | 0.000 00 | 0.000 00 |
| Towns/Suburbs | Univariable | −0.004 95 | −0.004 73 | −0.012 84 | 0.001 52 |
| Cities | Univariable | 0.011 90 | 0.011 94 | −0.002 94 | 0.026 40 |
| Fragility = 1 (Low) | Univariable | 0.000 00 | 0.000 00 | 0.000 00 | 0.000 00 |
| Fragility = 2 | Univariable | **0.019 70** | **0.019 78** | **0.011 81** | **0.027 28** |
| Fragility = 3 | Univariable | **0.027 81** | **0.028 09** | **0.019 00** | **0.035 26** |
| Fragility = 4 | Univariable | **0.034 56** | **0.034 81** | **0.024 98** | **0.043 47** |
| Fragility = 5 (High) | Univariable | **0.030 15** | **0.030 37** | **0.019 05** | **0.041 15** |
| **Multivariable model** | | | | | |
| Green Spaces | Multivariable | **−0.005 61** | **−0.005 56** | **−0.009 69** | **−0.001 14** |
| Average Temperature | Multivariable | 0.000 31 | 0.000 47 | −0.006 24 | 0.007 66 |
| Hospital Beds | Multivariable | −0.002 94 | −0.002 90 | −0.006 74 | 0.000 73 |
| Mid Population Density (ref.) | Multivariable | 0.000 00 | 0.000 00 | 0.000 00 | 0.000 00 |
| Low Population Density | Multivariable | −0.005 51 | −0.005 55 | −0.011 93 | 0.000 89 |
| High Population Density | Multivariable | −0.003 16 | −0.003 32 | −0.010 64 | 0.004 44 |
| Proportion 85+ | Multivariable | **0.007 15** | **0.007 05** | **0.003 79** | **0.010 64** |
| Health Expenditure | Multivariable | **0.008 39** | **0.008 39** | **0.004 04** | **0.013 07** |
| Smoking | Multivariable | 0.003 68 | 0.003 70 | −0.002 33 | 0.009 43 |
| Obesity | Multivariable | 0.000 55 | 0.000 40 | −0.004 65 | 0.005 83 |
| Rural Areas (ref.) | Multivariable | 0.000 00 | 0.000 00 | 0.000 00 | 0.000 00 |
| Towns/Suburbs | Multivariable | 0.005 70 | 0.005 55 | −0.001 12 | 0.012 56 |
| Cities | Multivariable | **0.017 91** | **0.017 70** | **0.004 29** | **0.031 90** |
| Fragility = 1 (Low) | Multivariable | 0.000 00 | 0.000 00 | 0.000 00 | 0.000 00 |
| Fragility = 2 | Multivariable | **0.015 94** | **0.015 81** | **0.009 25** | **0.022 96** |
| Fragility = 3 | Multivariable | **0.022 88** | **0.022 83** | **0.014 71** | **0.030 15** |
| Fragility = 4 | Multivariable | **0.027 18** | **0.027 39** | **0.018 70** | **0.035 69** |
| Fragility = 5 (High) | Multivariable | **0.020 05** | **0.019 77** | **0.009 29** | **0.030 97** |

**Table S8:** Association estimates of ERH considering the single components of the Fragility Index from univariable, multivariable, and spatial effect modification models

| Variable | Model | Mean | Median | 95% CrI lower | 95% CrI upper |
|---|---|---|---|---|---|
| **Dependency Index** | | | | | |
| Q1 (Low) | Univariable | 0.00 | 0.00 | 0.00 | 0.00 |
| Q2 | Univariable | 0.08 | 0.07 | -2.72 | 3.11 |
| Q3 | Univariable | 2.65 | 2.71 | -0.68 | 5.79 |
| Q4 | Univariable | **3.33** | **3.32** | **0.05** | **6.43** |
| Q5 (High) | Univariable | **4.51** | **4.46** | **1.19** | **7.75** |
| Q1 (Low) | Multivariable | 0.00 | 0.00 | 0.00 | 0.00 |
| Q2 | Multivariable | -0.56 | -0.57 | -2.86 | 1.76 |
| Q3 | Multivariable | 1.36 | 1.33 | -1.19 | 4.33 |
| Q4 | Multivariable | 1.95 | 1.96 | -0.96 | 4.86 |
| Q5 (High) | Multivariable | **3.27** | **3.23** | **0.00** | **6.77** |
| Q1 (Low) | Spatial | 0.00 | 0.00 | 0.00 | 0.00 |
| Q2 | Spatial | 0.50 | 0.47 | -1.20 | 2.32 |
| Q3 | Spatial | 1.03 | 1.04 | -0.87 | 2.83 |
| Q4 | Spatial | 0.36 | 0.37 | -1.67 | 2.28 |
| Q5 (High) | Spatial | 0.32 | 0.33 | -2.16 | 2.74 |
| **Employment Rate** | | | | | |
| Q1 (Low) | Univariable | 0.00 | 0.00 | 0.00 | 0.00 |
| Q2 | Univariable | **-12.73** | **-12.72** | **-16.25** | **-9.14** |
| Q3 | Univariable | **-26.06** | **-25.98** | **-31.10** | **-21.62** |
| Q4 | Univariable | **-32.16** | **-32.07** | **-37.01** | **-27.73** |
| Q5 (High) | Univariable | **-37.94** | **-37.88** | **-42.76** | **-33.71** |
| Q1 (Low) | Multivariable | 0.00 | 0.00 | 0.00 | 0.00 |
| Q2 | Multivariable | **-4.15** | **-4.11** | **-7.49** | **-0.95** |
| Q3 | Multivariable | -3.77 | -3.75 | -7.86 | 0.20 |
| Q4 | Multivariable | **-10.86** | **-10.81** | **-15.22** | **-6.66** |
| Q5 (High) | Multivariable | **-10.79** | **-10.82** | **-15.21** | **-6.29** |
| Q1 (Low) | Spatial | 0.00 | 0.00 | 0.00 | 0.00 |
| Q2 | Spatial | **-3.08** | **-3.07** | **-5.43** | **-0.91** |
| Q3 | Spatial | -2.24 | -2.21 | -5.35 | 0.54 |
| Q4 | Spatial | **-3.32** | **-3.29** | **-6.73** | **-0.02** |
| Q5 (High) | Spatial | **-4.23** | **-4.22** | **-7.67** | **-0.98** |
| **Firms per Capita** | | | | | |
| Q1 (Low) | Univariable | 0.00 | 0.00 | 0.00 | 0.00 |
| Q2 | Univariable | -2.67 | -2.65 | -5.51 | 0.03 |
| Q3 | Univariable | **-8.36** | **-8.38** | **-11.50** | **-5.21** |
| Q4 | Univariable | **-13.35** | **-13.34** | **-16.78** | **-10.32** |
| Q5 (High) | Univariable | **-19.14** | **-19.09** | **-22.72** | **-15.90** |
| Q1 (Low) | Multivariable | 0.00 | 0.00 | 0.00 | 0.00 |
| Q2 | Multivariable | -0.59 | -0.59 | -2.82 | 1.58 |
| Q3 | Multivariable | **-2.72** | **-2.67** | **-5.22** | **-0.17** |
| Q4 | Multivariable | **-4.62** | **-4.62** | **-7.28** | **-2.08** |
| Q5 (High) | Multivariable | **-4.93** | **-4.99** | **-7.64** | **-2.01** |
| Q1 (Low) | Spatial | 0.00 | 0.00 | 0.00 | 0.00 |
| Q2 | Spatial | -0.74 | -0.72 | -2.62 | 1.00 |
| Q3 | Spatial | -1.53 | -1.53 | -3.45 | 0.31 |
| Q4 | Spatial | -1.74 | -1.70 | -3.76 | 0.29 |
| Q5 (High) | Spatial | **-2.84** | **-2.81** | **-5.03** | **-0.67** |
| **Land Consumption** | | | | | |

| Variable | Model | Mean | Median | 95% CrI lower | 95% CrI upper |
|---|---|---|---|---|---|
| Q1 (Low) | Univariable | 0.00 | 0.00 | 0.00 | 0.00 |
| Q2 | Univariable | **13.39** | **13.36** | **10.27** | **16.43** |
| Q3 | Univariable | **15.74** | **15.71** | **12.41** | **19.18** |
| Q4 | Univariable | **10.47** | **10.54** | **6.56** | **14.02** |
| Q5 (High) | Univariable | 1.76 | 1.84 | -1.91 | 5.34 |
| Q1 (Low) | Multivariable | 0.00 | 0.00 | 0.00 | 0.00 |
| Q2 | Multivariable | 2.15 | 2.16 | -1.13 | 5.12 |
| Q3 | Multivariable | 0.09 | 0.07 | -3.66 | 3.84 |
| Q4 | Multivariable | **-8.42** | **-8.45** | **-12.84** | **-3.94** |
| Q5 (High) | Multivariable | **-19.22** | **-19.25** | **-24.37** | **-13.71** |
| Q1 (Low) | Spatial | 0.00 | 0.00 | 0.00 | 0.00 |
| Q2 | Spatial | 0.47 | 0.49 | -1.97 | 2.84 |
| Q3 | Spatial | -0.29 | -0.30 | -3.02 | 2.65 |
| Q4 | Spatial | -2.96 | -2.85 | -6.47 | 0.24 |
| Q5 (High) | Spatial | **-6.41** | **-6.15** | **-16.28** | **-2.33** |
| **Landslide Risk** | | | | | |
| Q1 (Low) | Univariable | 0.00 | 0.00 | 0.00 | 0.00 |
| Q2 | Univariable | **14.66** | **14.68** | **11.04** | **18.48** |
| Q3 | Univariable | **13.07** | **13.04** | **8.89** | **17.47** |
| Q4 | Univariable | **9.52** | **9.49** | **5.24** | **13.72** |
| Q5 (High) | Univariable | **5.03** | **5.03** | **0.59** | **9.30** |
| Q1 (Low) | Multivariable | 0.00 | 0.00 | 0.00 | 0.00 |
| Q2 | Multivariable | **3.03** | **3.01** | **0.20** | **6.17** |
| Q3 | Multivariable | **10.13** | **10.17** | **6.49** | **13.62** |
| Q4 | Multivariable | **10.39** | **10.34** | **6.61** | **14.30** |
| Q5 (High) | Multivariable | **9.62** | **9.63** | **5.61** | **13.52** |
| Q1 (Low) | Spatial | 0.00 | 0.00 | 0.00 | 0.00 |
| Q2 | Spatial | 0.20 | 0.18 | -2.23 | 2.99 |
| Q3 | Spatial | 1.83 | 1.77 | -0.80 | 4.61 |
| Q4 | Spatial | 1.74 | 1.62 | -1.05 | 4.81 |
| Q5 (High) | Spatial | 1.89 | 1.80 | -1.05 | 5.11 |
| **Low Education Rate** | | | | | |
| Q1 (Low) | Univariable | 0.00 | 0.00 | 0.00 | 0.00 |
| Q2 | Univariable | **9.18** | **9.15** | **6.56** | **12.09** |
| Q3 | Univariable | **13.03** | **13.02** | **10.19** | **16.02** |
| Q4 | Univariable | **16.25** | **16.26** | **13.00** | **19.47** |
| Q5 (High) | Univariable | **20.51** | **20.43** | **17.06** | **23.69** |
| Q1 (Low) | Multivariable | 0.00 | 0.00 | 0.00 | 0.00 |
| Q2 | Multivariable | **3.35** | **3.30** | **1.21** | **5.65** |
| Q3 | Multivariable | **3.91** | **3.93** | **1.49** | **6.55** |
| Q4 | Multivariable | **4.98** | **5.00** | **2.34** | **7.61** |
| Q5 (High) | Multivariable | 2.15 | 2.10 | -0.97 | 5.29 |
| Q1 (Low) | Spatial | 0.00 | 0.00 | 0.00 | 0.00 |
| Q2 | Spatial | **1.97** | **2.01** | **0.14** | **3.77** |
| Q3 | Spatial | **3.03** | **3.06** | **0.98** | **4.92** |
| Q4 | Spatial | **3.21** | **3.20** | **1.11** | **5.23** |
| Q5 (High) | Spatial | **3.05** | **3.05** | **0.87** | **5.32** |
| **Low Productivity Employees** | | | | | |
| Q1 (Low) | Univariable | 0.00 | 0.00 | 0.00 | 0.00 |
| Q2 | Univariable | **8.45** | **8.45** | **5.93** | **10.80** |
| Q3 | Univariable | **17.22** | **17.23** | **14.51** | **20.17** |
| Q4 | Univariable | **26.89** | **26.92** | **23.72** | **30.05** |

| Variable | Model | Mean | Median | 95% CrI lower | 95% CrI upper |
|---|---|---|---|---|---|
| Q5 (High) | Univariable | **29.45** | **29.34** | **25.52** | **33.41** |
| Q1 (Low) | Multivariable | 0.00 | 0.00 | 0.00 | 0.00 |
| Q2 | Multivariable | **2.86** | **2.87** | **0.80** | **4.99** |
| Q3 | Multivariable | **6.73** | **6.65** | **4.41** | **9.28** |
| Q4 | Multivariable | **9.88** | **9.79** | **7.20** | **12.82** |
| Q5 (High) | Multivariable | **10.95** | **10.87** | **7.89** | **14.18** |
| Q1 (Low) | Spatial | 0.00 | 0.00 | 0.00 | 0.00 |
| Q2 | Spatial | 0.83 | 0.84 | -0.81 | 2.52 |
| Q3 | Spatial | 0.91 | 0.82 | -1.10 | 4.81 |
| Q4 | Spatial | 1.89 | 1.70 | -0.09 | 6.90 |
| Q5 (High) | Spatial | **2.46** | **2.19** | **0.06** | **8.71** |
| **Migration Rate** | | | | | |
| Q1 (Low) | Univariable | 0.00 | 0.00 | 0.00 | 0.00 |
| Q2 | Univariable | 0.53 | 0.57 | -2.30 | 3.18 |
| Q3 | Univariable | **-6.32** | **-6.30** | **-9.29** | **-3.15** |
| Q4 | Univariable | **-9.88** | **-9.83** | **-13.20** | **-6.61** |
| Q5 (High) | Univariable | **-9.09** | **-9.10** | **-12.51** | **-5.69** |
| Q1 (Low) | Multivariable | 0.00 | 0.00 | 0.00 | 0.00 |
| Q2 | Multivariable | 1.46 | 1.45 | -0.80 | 3.66 |
| Q3 | Multivariable | -0.28 | -0.34 | -2.59 | 2.03 |
| Q4 | Multivariable | -1.15 | -1.17 | -3.61 | 1.52 |
| Q5 (High) | Multivariable | 1.54 | 1.56 | -0.93 | 4.22 |
| Q1 (Low) | Spatial | 0.00 | 0.00 | 0.00 | 0.00 |
| Q2 | Spatial | 1.49 | 1.52 | -0.29 | 3.11 |
| Q3 | Spatial | 1.14 | 1.13 | -0.74 | 2.95 |
| Q4 | Spatial | 0.68 | 0.67 | -1.20 | 2.53 |
| Q5 (High) | Spatial | **2.03** | **1.99** | **0.02** | **3.98** |
| **Protected Natural Areas (%)** | | | | | |
| Q1 (Low) | Univariable | 0.00 | 0.00 | 0.00 | 0.00 |
| Q2 | Univariable | **16.94** | **16.98** | **12.69** | **21.30** |
| Q3 | Univariable | **8.46** | **8.50** | **4.99** | **11.93** |
| Q4 | Univariable | **8.02** | **8.02** | **4.45** | **11.58** |
| Q5 (High) | Univariable | 1.29 | 1.19 | -2.54 | 5.20 |
| Q1 (Low) | Multivariable | 0.00 | 0.00 | 0.00 | 0.00 |
| Q2 | Multivariable | -3.11 | -3.14 | -6.81 | 0.60 |
| Q3 | Multivariable | -2.87 | -2.82 | -5.99 | 0.15 |
| Q4 | Multivariable | -2.43 | -2.42 | -5.39 | 0.93 |
| Q5 (High) | Multivariable | -2.80 | -2.82 | -5.70 | 0.21 |
| Q1 (Low) | Spatial | 0.00 | 0.00 | 0.00 | 0.00 |
| Q2 | Spatial | 2.16 | 2.22 | -0.44 | 4.76 |
| Q3 | Spatial | 1.37 | 1.39 | -0.82 | 3.49 |
| Q4 | Spatial | 0.63 | 0.66 | -1.56 | 2.82 |
| Q5 (High) | Spatial | -0.42 | -0.46 | -2.67 | 1.79 |
| **High Transport Emission Rate** | | | | | |
| Q1 (Low) | Univariable | 0.00 | 0.00 | 0.00 | 0.00 |
| Q2 | Univariable | **7.73** | **7.67** | **4.82** | **10.86** |
| Q3 | Univariable | **18.88** | **18.85** | **15.91** | **22.11** |
| Q4 | Univariable | **32.18** | **32.09** | **28.69** | **35.82** |
| Q5 (High) | Univariable | **33.84** | **33.89** | **29.98** | **37.45** |
| Q1 (Low) | Multivariable | 0.00 | 0.00 | 0.00 | 0.00 |
| Q2 | Multivariable | 1.54 | 1.54 | -0.96 | 4.03 |
| Q3 | Multivariable | **7.60** | **7.55** | **4.88** | **10.57** |

| Variable | Model | Mean | Median | 95% CrI lower | 95% CrI upper |
|---|---|---|---|---|---|
| Q4 | Multivariable | **12.88** | **12.87** | **9.70** | **16.55** |
| Q5 (High) | Multivariable | **11.95** | **11.90** | **7.95** | **16.23** |
| Q1 (Low) | Spatial | 0.00 | 0.00 | 0.00 | 0.00 |
| Q2 | Spatial | 0.41 | 0.41 | -1.66 | 2.52 |
| Q3 | Spatial | 1.35 | 1.18 | -1.10 | 5.65 |
| Q4 | Spatial | 2.79 | 2.54 | -0.20 | 10.12 |
| Q5 (High) | Spatial | **3.60** | **3.43** | **0.46** | **8.59** |
| **Time To Access Services** | | | | | |
| Q1 (Low) | Univariable | 0.00 | 0.00 | 0.00 | 0.00 |
| Q2 | Univariable | **5.24** | **5.30** | **2.42** | **7.91** |
| Q3 | Univariable | **8.03** | **7.98** | **5.18** | **10.91** |
| Q4 | Univariable | **8.63** | **8.64** | **5.38** | **11.93** |
| Q5 (High) | Univariable | 0.38 | 0.33 | -3.45 | 4.08 |
| Q1 (Low) | Multivariable | 0.00 | 0.00 | 0.00 | 0.00 |
| Q2 | Multivariable | 2.20 | 2.20 | -0.10 | 4.62 |
| Q3 | Multivariable | **4.06** | **4.08** | **1.49** | **6.82** |
| Q4 | Multivariable | 2.84 | 2.84 | -0.12 | 5.97 |
| Q5 (High) | Multivariable | 0.82 | 0.73 | -2.85 | 4.60 |
| Q1 (Low) | Spatial | 0.00 | 0.00 | 0.00 | 0.00 |
| Q2 | Spatial | 1.06 | 1.04 | -0.91 | 3.07 |
| Q3 | Spatial | **2.25** | **2.24** | **0.01** | **4.62** |
| Q4 | Spatial | 1.53 | 1.52 | -1.03 | 4.11 |
| Q5 (High) | Spatial | 0.98 | 1.02 | -2.42 | 4.07 |
| **Unsorted Waste** | | | | | |
| Q1 (Low) | Univariable | 0.00 | 0.00 | 0.00 | 0.00 |
| Q2 | Univariable | **7.82** | **7.81** | **4.84** | **10.79** |
| Q3 | Univariable | **9.50** | **9.51** | **6.43** | **12.58** |
| Q4 | Univariable | **8.14** | **8.02** | **4.93** | **11.52** |
| Q5 (High) | Univariable | **10.41** | **10.35** | **6.70** | **13.81** |
| Q1 (Low) | Multivariable | 0.00 | 0.00 | 0.00 | 0.00 |
| Q2 | Multivariable | **6.45** | **6.41** | **4.08** | **8.76** |
| Q3 | Multivariable | **11.10** | **11.10** | **8.64** | **13.60** |
| Q4 | Multivariable | **10.92** | **10.90** | **8.15** | **13.68** |
| Q5 (High) | Multivariable | **8.11** | **8.14** | **4.92** | **11.28** |
| Q1 (Low) | Spatial | 0.00 | 0.00 | 0.00 | 0.00 |
| Q2 | Spatial | 1.84 | 1.73 | -0.09 | 4.18 |
| Q3 | Spatial | 2.03 | 1.82 | -0.01 | 4.68 |
| Q4 | Spatial | **2.48** | **2.29** | **0.30** | **5.25** |
| Q5 (High) | Spatial | 0.97 | 0.82 | -1.51 | 4.28 |

**Figure S8:** Sensitivity analysis including the interaction terms. Median of the posterior distribution of the relative mortality risk across the 7,895 municipalities during 2011-2023 by temperature percentiles. The green line represents the nationwide relative mortality risk calculated as the median of the municipality-specific median relative mortality risk per percentile.

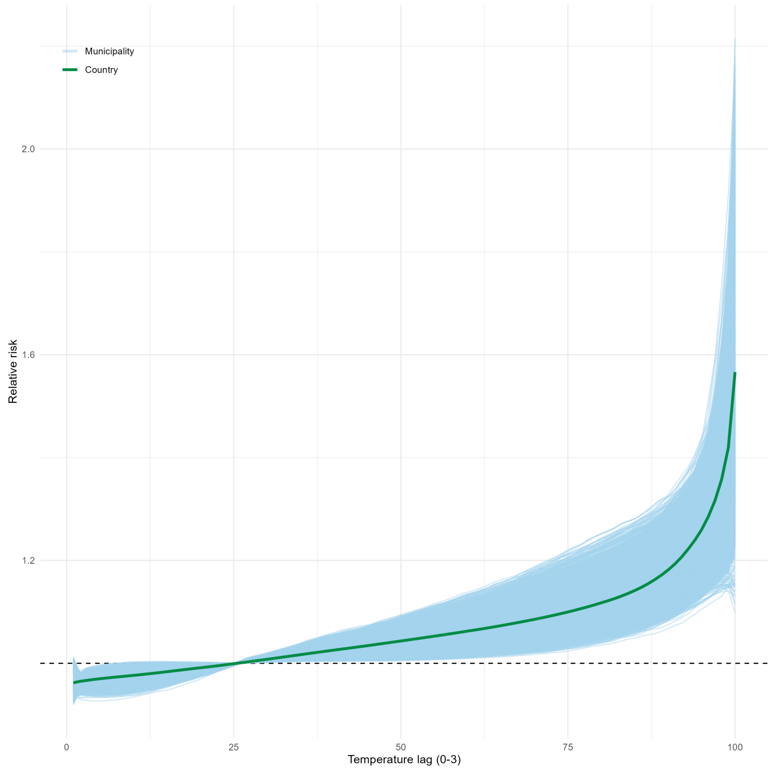

**Figure S9:** Sensitivity analysis removing the years after the start of the COVID-19 pandemic in Italy. Median of the posterior distribution of the relative mortality risk across the 7,895 municipalities during 2011-2019 by temperature percentiles. The green line represents the nationwide relative mortality risk calculated as the median of the municipality-specific median relative mortality risk per percentile.

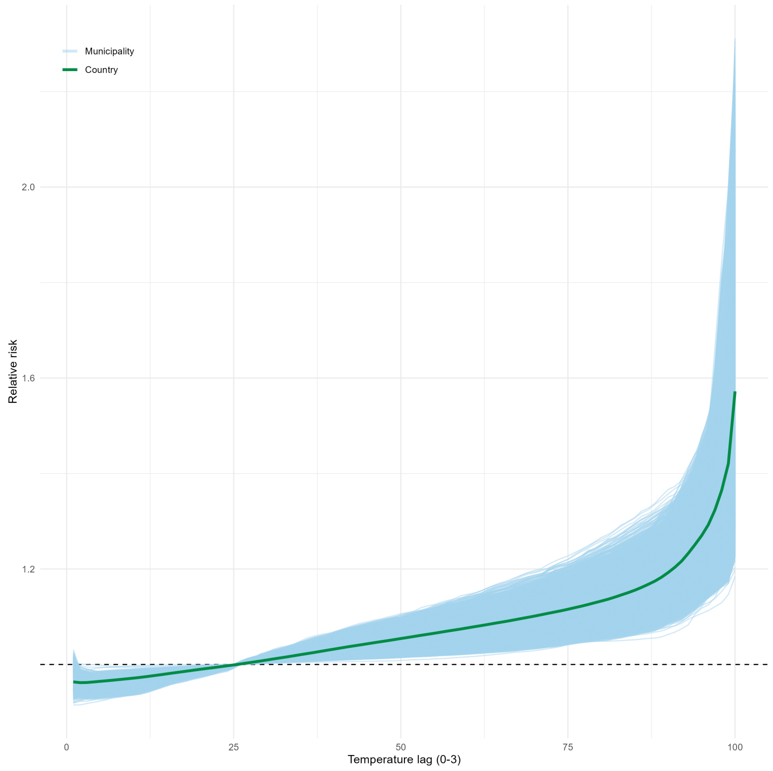

**Figure S10:** A. Median of the posterior distribution of the relative mortality risk at 95th temperature percentile at municipality level; B.Median of the posterior distribution of the relative mortality risk at 99th temperature percentile at municipality level; C. The exceedance probability of relative mortality risk at 95th temperature percentile in the area is higher than the mean value of the RR at 95th percentile across the country; D. The exceedance probability of relative mortality risk at 99th temperature percentile in the area is higher than the mean value of the RR at 95th percentile across the country

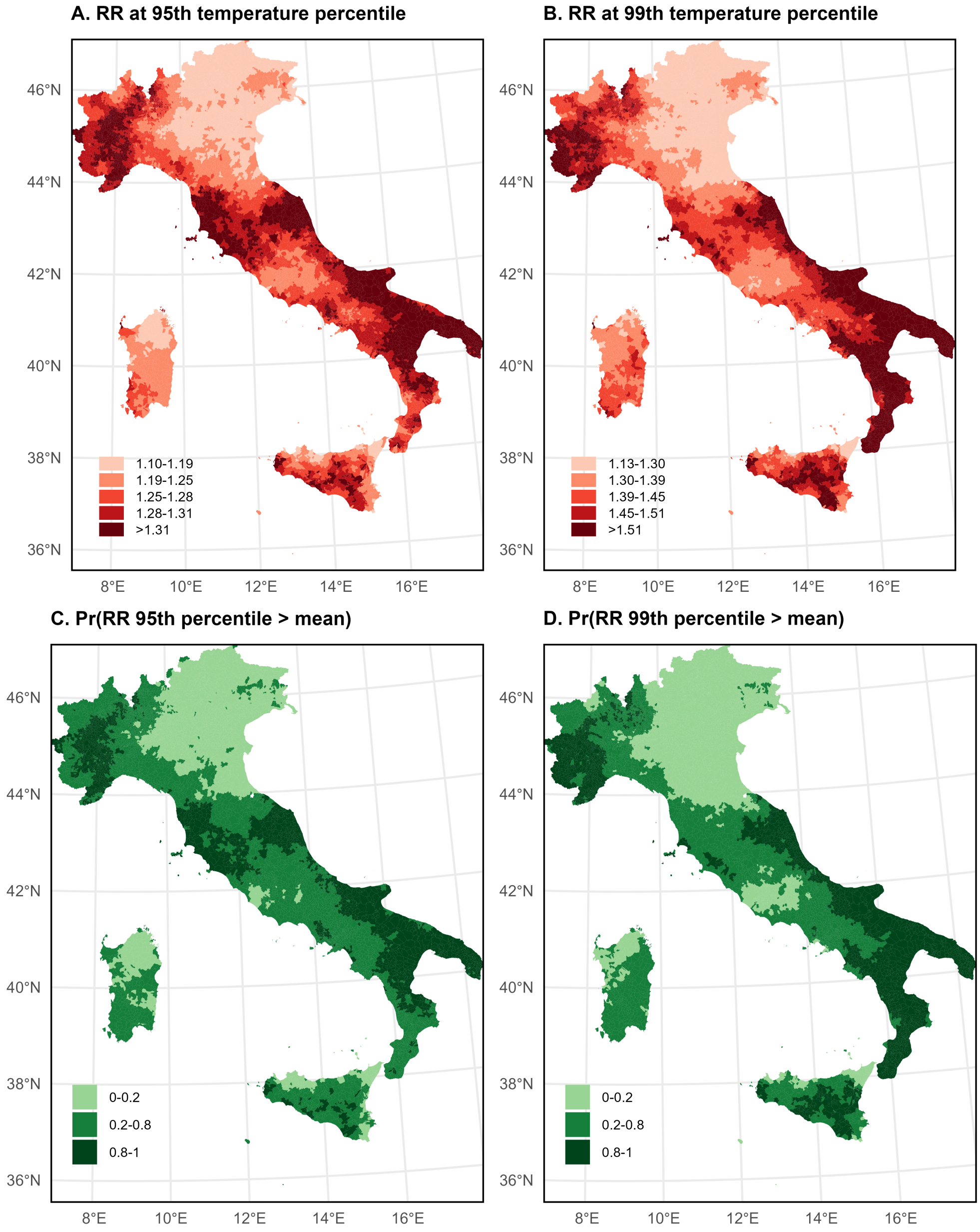

### Text

#### Text S1: Population

We assumed that the population remained constant over the summer period within each year, as short-term variations due to births and deaths can be negligible, particularly in smaller municipalities. Due to municipal mergers and splits, the number of municipalities included in the population data varied across years. To ensure compatibility with the mortality data recorded in 2023, we harmonised the population datasets by aligning them with the municipal structure of that year.

#### Text S2: Community factors

We first considered factors related with environmental conditions. We measured green space coverage using the Normalized Difference Vegetation Index (NDVI) from ERA-5 and converted the monthly data into a spatial-only metric by averaging values during the study period by municipality. We included green space because it may influence local microclimates, modify heat exposure [1], and affect vulnerability through its association with chronic conditions. We also used average temperature from ERA-5 as an indicator of local adaptation to heat, computing a spatial-only metric by averaging temperature values over the study period for each municipality.

To assess the role of demographic factors, we included population density (ISTAT), urbanicity (EUROSTAT), and the proportion of individuals aged 85 years or older (ISTAT). Urban environments may influence heat exposure and vulnerability because of higher temperatures associated with the urban heat island effect, as well as differences in the built environment [1, 2]. We also included the proportion of individuals aged 85 years or older calculated as the average proportion during the study period by municipality, given their heightened vulnerability to heat-related health impacts.

We also estimated the impact of health indicators, including number of hospital beds per population, health expenditure per population, and smoking and obesity prevalence. Hospital bed availability and health expenditure were included as proxies for healthcare system capacity and access to medical resources across areas. These variables were obtained from the Italian Ministry of Health at the provincial level for the year 2018 and were included as the number of hospital beds per unit and expenditure on medical devices per unit, respectively. Smoking and obesity prevalence were selected as proxies for the overall health status of the population in each municipality, as a higher burden of chronic conditions is associated with increased vulnerability to heat-related mortality. Obesity prevalence was defined as the proportion of individuals aged 18 years and older who were classified as obese in 2023 at municipal level (ISTAT). Similarly, smoking prevalence was defined as the proportion of individuals aged 14 years and older who reported being smokers in 2023 at municipal level (ISTAT).

#### Text S3: Priors

We specified prior distributions on all parameters in the model. For the fixed effects, $\beta_0, \boldsymbol{\beta}, \boldsymbol{\gamma}$, we used flat, non-informative priors. We used conditional autoregressive priors for the spatially, $\boldsymbol{\beta}'_m$ and $b_m$, and temporally, $\omega_d$ and $\delta_t$, varying parameters. These priors allow adjacent municipalities and successive days and years to be more similar than non-adjacent locations or time points [3]. Briefly, we used a re-parameterised Besag-York-Mollié (BYM2) prior [4, 3] for the spatially varying random effects, a random walk of order 2 to model seasonality and an iid (independent and identically distributed) component to model long-term trends. We use Penalised Complexity (PC) priors for hyper-parameters in the prior distributions as in previous work [4].

#### Text S4: Epidemiological metric

From the first-stage model we define heat-related risks as the posterior distribution of the RR at the 90th temperature percentile of each municipality, the attributable fraction due to heat (AFH) and the number of excess deaths due to heat: excess risk due to heat (ERH). All estimates were relative to the posterior distribution of the municipality-specific Minimum Mortality Temperature (MMT), i.e. the distribution of temperatures at which the lowest mortality risk is observed [5]. In line with the literature we restricted the MMT to the 25th–90th percentile range of summer temperatures. As we are interested in the effect of heat, we calculate AFH and ERH for temperatures higher than the MMT. As we are interested in the effect of heat, we calculated the heat-attributable fraction (AFH) and the heat-attributable excess mortality rate (ERH) for temperatures above the minimum mortality temperature (MMT). The heat-attributable fraction for each municipality ($m$) quantifies the proportion of summer mortality attributable to heat exposure and was estimated as:

$$AFH_m(x) = \frac{RR_m(x) - 1}{RR_m(x)}, \tag{1}$$

where $RR_m(x)$ denotes the relative risk associated with temperature $x$ in municipality $m$.

We then estimated the heat-attributable excess mortality rate (ERH), defined as the number of heat-attributable deaths per population:

$$ERH_m = \frac{Y'_m}{P_m} = \frac{AFH_m Y_m}{P_m}, \tag{2}$$

where $Y'_m$ denotes the estimated number of deaths attributable to heat exposure, $Y_m$ is the total number of deaths, and $P_m$ is the population of municipality $m$.
